# *Mycobacterium tuberculosis*-dependent Monocyte Expression Quantitative Trait Loci and Tuberculosis Pathogenesis

**DOI:** 10.1101/2023.08.28.23294698

**Authors:** Hyejeong Hong, Kimberly A. Dill-McFarland, Jason D. Simmons, Glenna J. Peterson, Penelope Benchek, Harriet Mayanja-Kizza, W. Henry Boom, Catherine M. Stein, Thomas R. Hawn

**Affiliations:** Department of Biobehavioral Health Sciences, School of Nursing, University of Pennsylvania, Philadelphia, PA, USA; Department of Medicine, University of Washington, Seattle, WA, USA; Department of Population & Quantitative Health Sciences, Case Western Reserve University, Cleveland, OH, USA; Department of Medicine, School of Medicine, Makerere University, Kampala, Uganda; Department of Medicine, Case Western Reserve University, Cleveland, OH, USA

**Author notes:** Corresponding author: Hyejeong Hong, University of Pennsylvania, School of Nursing, Department of Biobehavioral Health Sciences, Claire M. Fagin Hall, 418 Curie Boulevard, Room 335, Philadelphia, PA 19104 – 4217, USA.

**Keywords:** Mycobacterium tuberculosis, expression quantitative trait loci, monocyte response, pro-inflammatory cytokine

## Abstract

The heterogeneity of outcomes after *Mycobacterium tuberculosis* (Mtb) exposure is a conundrum associated with millennia of host-pathogen co-evolution. We hypothesized that human myeloid cells contain genetically encoded, Mtb-specific responses that regulate critical steps in tuberculosis (TB) pathogenesis. We mapped genome-wide expression quantitative trait loci (eQTLs) in Mtb-infected monocytes with RNAseq from 80 Ugandan household contacts of pulmonary TB cases to identify monocyte-specific, Mtb-dependent eQTLs and their association with cytokine expression and clinical resistance to tuberculin skin test (TST) and interferon-γ release assay (IGRA) conversion. cis-eQTLs (n=1,567) were identified in Mtb-infected monocytes (FDR<0.01), including 29 eQTLs in 16 genes which were Mtb-dependent (significant for Mtb:genotype interaction [FDR<0.1], but not classified as eQTL in media condition [FDR≥0.01]). A subset of eQTLs were associated with Mtb-induced cytokine expression (n=8) and/or clinical resistance to TST/IGRA conversion (n=1). Expression of *BMP6*, an Mtb-dependent eQTL gene, was associated with *IFNB1* induction in Mtb-infected and DNA ligand-induced cells. Network and enrichment analyses identified fatty acid metabolism as a pathway associated with eQTL genes. These findings suggest that monocyte genes contain Mtb-dependent eQTLs, including a subset associated with cytokine expression and/or clinical resistance to TST/IGRA conversion, providing insight into immunogenetic pathways regulating susceptibility to Mtb infection and TB pathogenesis.

## INTRODUCTION

Despite ongoing efforts to eradicate tuberculosis (TB), it remains one of the leading infectious diseases worldwide [1]. The wide spectrum of clinical states, ranging from asymptomatic latent TB infection (LTBI) to active TB disease, presents a challenge in identifying immune events providing protection early after exposure. While various inter-individual and environmental factors can influence TB susceptibility, such as bacillary load, proximity, duration of contact, age, malnutrition, and hygienic conditions, host genetics and immunologic factors are believed to play a significant role in increasing risk for TB disease [2, 3]. Therefore, it is critical to identify immunogenetic determinants of TB susceptibility to develop effective prevention strategies and provide insights for new TB treatments.

*Mycobacterium tuberculosis* (Mtb), the primary causative pathogen of TB, is an ancient infectious agent and has co-evolved with humans for over 10,000 years [4]. Host and pathogen genetic pressure over this long time period selects genes and variants that may regulate protective immune responses. The primary niche of this evolutionary battle occurs within myeloid cells where Mtb resides in a phagosome. Such pressures can contribute to inter-individual variation in susceptibility to Mtb infection and TB disease [5]; however, the specific major susceptibility genes and variants remain largely unknown. The search for the genetic determinants of clinical TB susceptibility has included case-control studies of candidate genes and genome-wide association studies (GWAS) [6–11]. Despite extensive efforts, the major genes and specific functional effects of the polymorphisms associated with TB remain poorly understood. One approach to address this knowledge gap is expression quantitative trait loci (eQTL) mapping [12]. By linking genetic variants with Mtb-dependent changes in intermediate traits within myeloid cells, such as cytokine gene expression, eQTL mapping can provide insights into underlying immunogenetic mechanisms of disease.

To investigate the functional consequences of genetic variants within macrophages responding to Mtb infection, we examine our hypothesis that a specific subset of monocyte genes has eQTLs that regulate pro-inflammatory responses to Mtb or that are associated with clinical TB phenotypes. Through genome-wide genotyping and transcriptional profiling of Mtb-infected and uninfected CD14+ monocytes from 80 household contacts (HHCs) of pulmonary TB in Uganda, we identified 29 eQTLs associated with the expression of 16 genes under Mtb-stimulated and not unstimulated conditions. Some of these ‘Mtb-dependent’ eQTLs were also associated with Mtb DNA sensor-dependent interferon-β (IFN-β) expression, resistance to tuberculin skin test (TST) and interferon-γ (IFN-γ) release assay (IGRA) conversion after Mtb exposure, and modulation of immunometabolic pathways. These findings shed light on the immunogenetic factors underlying TB susceptibility and host-pathogen interactions in the context of Mtb infection.

## RESULTS

### Monocytes express 16 genes with Mtb-dependent eQTLs

To determine whether monocyte responses to Mtb include stimulation-specific eQTLs, we examined genome-wide genotyping (MEGA^EX^ genotyping array) and RNAseq transcriptional profiles of cells infected with H37Rv for 6h or media controls in 101 participants in a previously described Ugandan TB household contact study [13–15]. We linked the genotypes and transcriptional profiles to define eQTLs among the 80 subjects from whom these datasets were available (Figure 1). Although this primary study compared individuals who did (LTBI) or did not (“resister” or RSTR) convert their TST/IGRA after heavy exposure, we did not include these phenotypes in our primary analysis. Both the RSTR and LTBI groups were relatively young (mean age = 23 years), with no significant differences observed in sex, body mass index (BMI), Bacille Calmette-Guérin (BCG) vaccination history, or epidemiologic exposure risk score between RSTR and LTBI groups (Table 1) [16]. To discover cis-eQTLs, we considered single nucleotide polymorphisms (SNPs) within 1 megabase (Mb) of the transcription start site (TSS) and their association with gene expression in the media and Mtb conditions. We observed 1,567 cis-eQTLs associated with the expression of 159 genes in Mtb-infected monocytes, whereas in uninfected monocytes, we identified 2,106 cis-eQTLs associated with the expression of 261 genes (false discovery rate [FDR] < 0.01, Table S1). There was a significant overlap of eQTLs identified in infected and uninfected monocytes (n = 1,269; Wilcoxon signed rank test, p < 0.001). To discover “Mtb-dependent” cis-eQTLs, we next evaluated the interaction effect of genotypes between Mtb-infected and uninfected monocytes, while controlling for age and sex (FDR < 0.1). Among the 1,567 eQTLs identified in Mtb-infected monocytes, we found 32 eQTLs associated with the expression of 17 genes that were significant for an Mtb-infection:genotype interaction (FDR <0.1). Three of these eQTL were also identified under unstimulated conditions (FDR <0.01), and thus were excluded (Figure 2a). For further analysis, we focused on the remaining 29 eQTLs, referred to as “Mtb-dependent eQTLs”, which were associated with the expression of 16 genes (Table S2, Figure S1).

**Figure 1.**
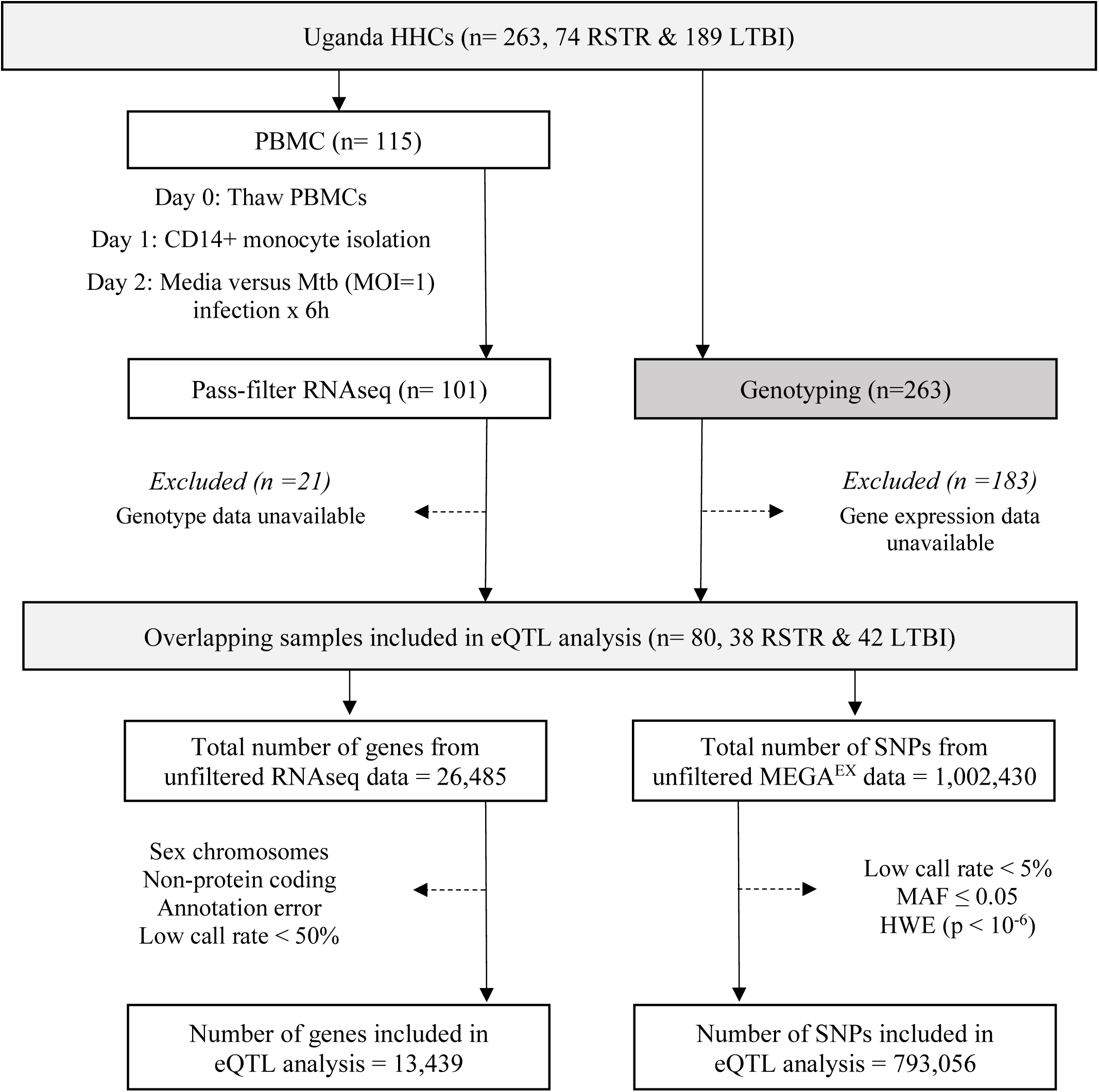
Study Design and eQTL Mapping Data Processing in the Uganda Household Contact Study. Household contacts (HHCs) of pulmonary TB index cases were studied, enrolled, and followed longitudinally for assessment using serial TST and IGRA testing. Laboratory studies included RNASeq measurements in Mtb-infected monocytes and genome-wide genotyping with the MEGA^EX^ genotyping array. eQTL = expression quantitative trait loci, HHC = household contact, HWE = Hardy-Weinberg equilibrium, IGRA = interferon-γ release assay, LTBI = latent tuberculosis infection, MAF = minor allele frequency, MOI = multiplicity of infection, Mtb = *Mycobacterium tuberculosis*, PBMC = peripheral blood mononuclear cell, RNAseq = RNA sequencing, RSTR = resister or resistant to TST/IGRA conversion after high TB exposure, SNP = single nucleotide polymorphism, TST = tuberculin skin test.

**Figure 2.**
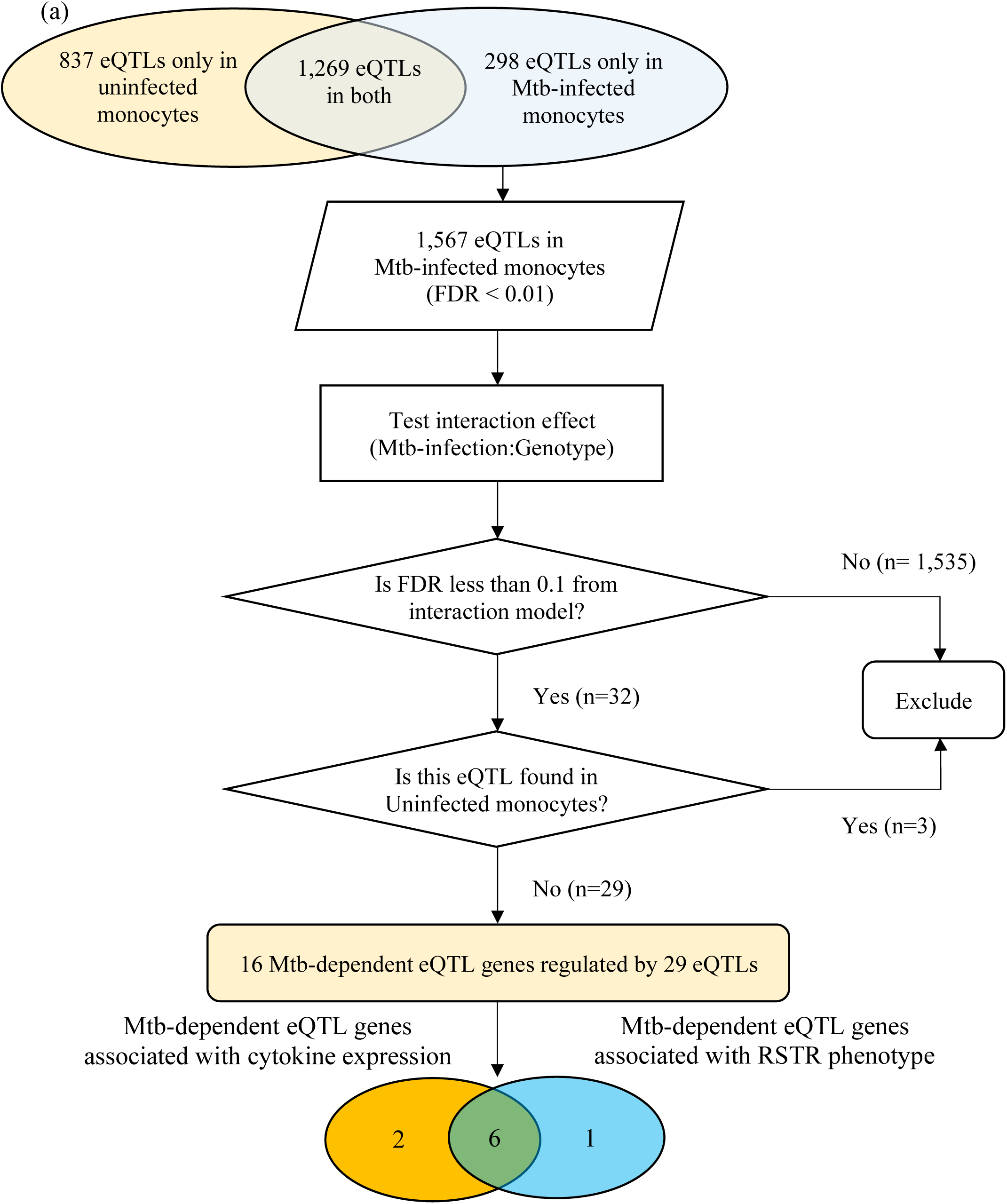
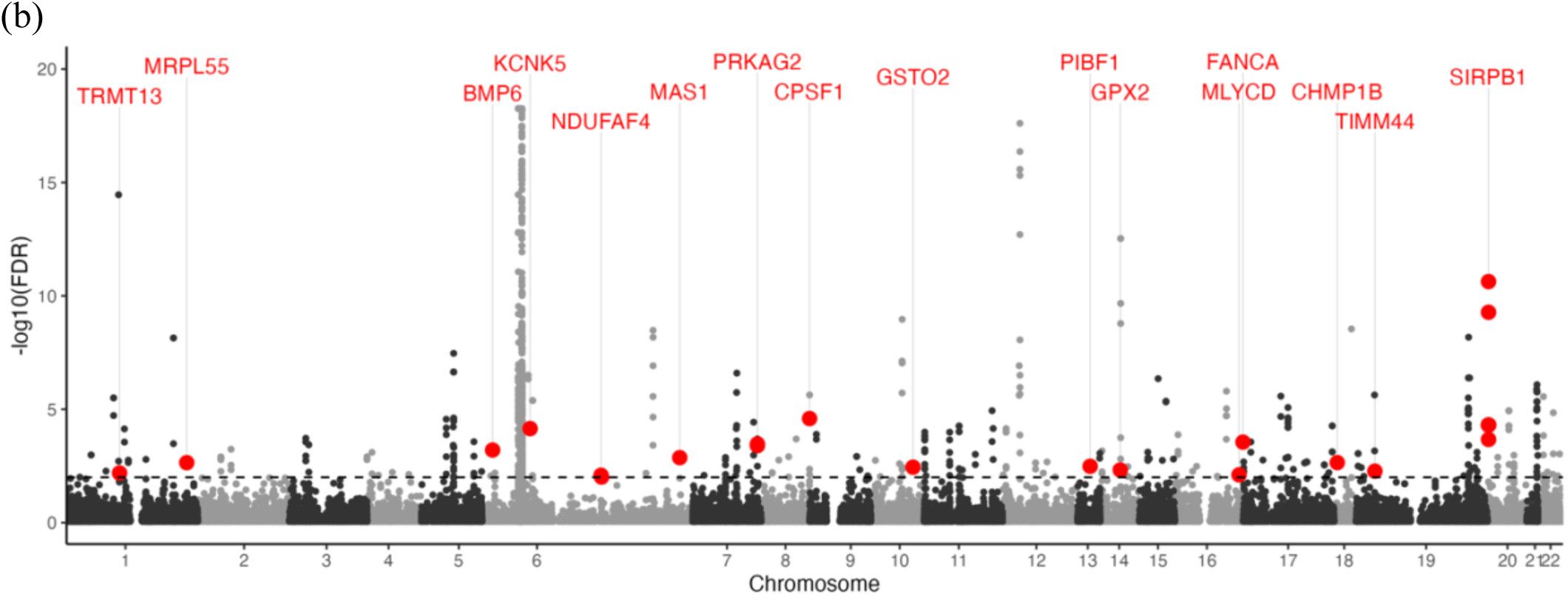
Mtb-dependent eQTL mapping: ***(a) Flowchart of Mtb-dependent eQTL identification***. Using an additive regression model (FDR< 0.01), we identified 2,106 eQTLs in uninfected monocytes and 1,567 eQTLs in Mtb-infected monocytes after adjusting for age and sex. Mtb-dependent eQTLs were specifically defined as cis-eQTLs that showed strong statistical evidence only in Mtb-infected monocytes. We performed additive linear regression with an interaction term for the Mtb stimulation status to evaluate the interaction effect at an FDR of 10%. Out of the 2,106 eQTLs in Mtb-infected monocytes, 32 eQTLs showed significant linear associations with gene expression levels after adjusting for age and sex (FDR <0.1). Three eQTLs were excluded as they were identified in both Mtb-infected and uninfected monocytes. We identified 29 Mtb-dependent eQTLs regulating the expression levels of 16 genes. Additionally, 8 Mtb-dependent eQTLs were associated with altered levels of TNF, IL6, IL1B, or IFNB1 expression *in trans* (orange circle). Finally, 7 Mtb-dependent eQTL genes contained SNPs associated with the clinical RSTR phenotype (blue circle). Importantly, 6 Mtb-dependent eQTL genes and variants were associated with both clinical RSTR phenotypes and the regulation of cytokine gene expression. ***(b) Manhattan plot of Mtb-dependent eQTLs.*** Significance of eQTLs in the Mtb-infected condition are plotted across the genome, and the horizontal dashed line represents the significance threshold (FDR = 0.01). Red dots mark lead eQTLs that are also significant in the interaction model (Mtb-infection:Genotype, FDR > 0.1), yet not in the media condition (FDR > 0.01); these are labeled with their respective cis genes. Grey dots positioned above this line represent eQTLs in Mtb-infected conditions (FDR < 0.01), though they do not demonstrate significance in the primary interaction model. eQTL = expression quantitative trait loci, FDR = false discovery rate, Mtb = *Mycobacterium tuberculosi*s, SNP = single nucleotide polymorphysm

**Table 1.**
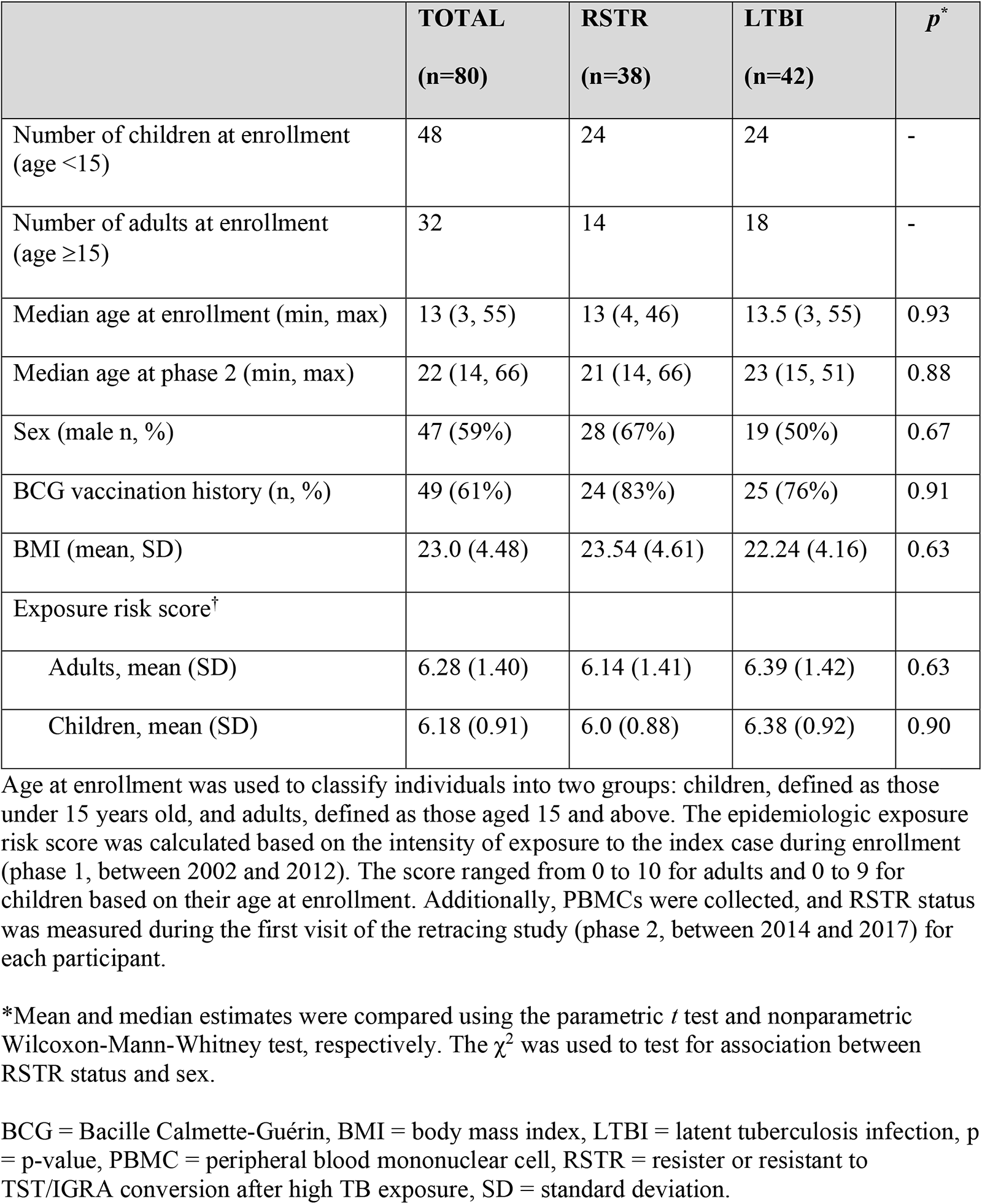
Demographic Characteristics of Participants.

|  | **TOTAL**<br>**(n=80)** | **RSTR**<br>**(n=38)** | **LTBI**<br>**(n=42)** | ***p*<sup>\*</sup>** |
| --- | --- | --- | --- | --- |
| Number of children at enrollment (age <15) | 48 | 24 | 24 | - |
| Number of adults at enrollment (age ≥15) | 32 | 14 | 18 | - |
| Median age at enrollment (min, max) | 13 (3, 55) | 13 (4, 46) | 13.5 (3, 55) | 0.93 |
| Median age at phase 2 (min, max) | 22 (14, 66) | 21 (14, 66) | 23 (15, 51) | 0.88 |
| Sex (male n, %) | 47 (59%) | 28 (67%) | 19 (50%) | 0.67 |
| BCG vaccination history (n, %) | 49 (61%) | 24 (83%) | 25 (76%) | 0.91 |
| BMI (mean, SD) | 23.0 (4.48) | 23.54 (4.61) | 22.24 (4.16) | 0.63 |
| Exposure risk score<sup>†</sup> |  |  |  |  |
| Adults, mean (SD) | 6.28 (1.40) | 6.14 (1.41) | 6.39 (1.42) | 0.63 |
| Children, mean (SD) | 6.18 (0.91) | 6.0 (0.88) | 6.38 (0.92) | 0.90 |

Age at enrollment was used to classify individuals into two groups: children, defined as those under 15 years old, and adults, defined as those aged 15 and above. The epidemiologic exposure risk score was calculated based on the intensity of exposure to the index case during enrollment (phase 1, between 2002 and 2012). The score ranged from 0 to 10 for adults and 0 to 9 for children based on their age at enrollment. Additionally, PBMCs were collected, and RSTR status was measured during the first visit of the retracing study (phase 2, between 2014 and 2017) for each participant.
\*Mean and median estimates were compared using the parametric *t* test and nonparametric Wilcoxon-Mann-Whitney test, respectively. The $\chi^2$ was used to test for association between RSTR status and sex.
BCG = Bacille Calmette-Guérin, BMI = body mass index, LTBI = latent tuberculosis infection, p = p-value, PBMC = peripheral blood mononuclear cell, RSTR = resister or resistant to TST/IGRA conversion after high TB exposure, SD = standard deviation.

Among the 16 Mtb-dependent eQTL genes (Table 2, Table S2, Figure 2b, Figure S1), 3 were associated with multiple eQTLs. Specifically, 12 eQTLs were associated with *SIRPB1* expression (Figure 3a), 2 eQTLs with *PRKAG2*, and 2 eQTLs with *NDUFAF4* (Figure S2). Notably, these eQTLs showed a high linkage disequilibrium (LD, r^2^ > 0.8), indicating a potential single causative locus for each gene. Mtb infection resulted in clear genotype-dependent effects as compared to the media condition. To further categorize the Mtb effects, we compared the slope change across genotypes that was observed under unstimulated and Mtb-infected conditions. Some genes (*CPSF1* and *SIRPB1)* had similar directionality of genotype effects in the media and Mtb condition that included a low FDR in the media condition (0.01 to 0.1), indicating that genetic effects are present even without Mtb infection, but they intensify following infection (Figure 3b,c). However, for the majority of the genes, the differences were more striking with higher FDR values in the media condition (FDR > 0.9) with low FDRs for the Mtb condition (< 0.01) (*BMP6, CHMP1B, FANCA, GSTO2, KCNK5, MAS1, MLYCD, NDUFAF4, PIBF1,* and *TRMT13*; Figure 3d,e and Figure S1). Finally, there were several genes where the directionality of genotype effect was different in the media and Mtb condition (*GPX2, MRPL55, PRKAG2, TIMM44*; Figure 3f,g, and Figure S1). In summary, our study identified 29 eQTLs that are associated with expression patterns of 16 genes in monocytes during Mtb infection, with genotype and condition-dependent patterns that vary in direction and magnitude.

**Figure 3.**
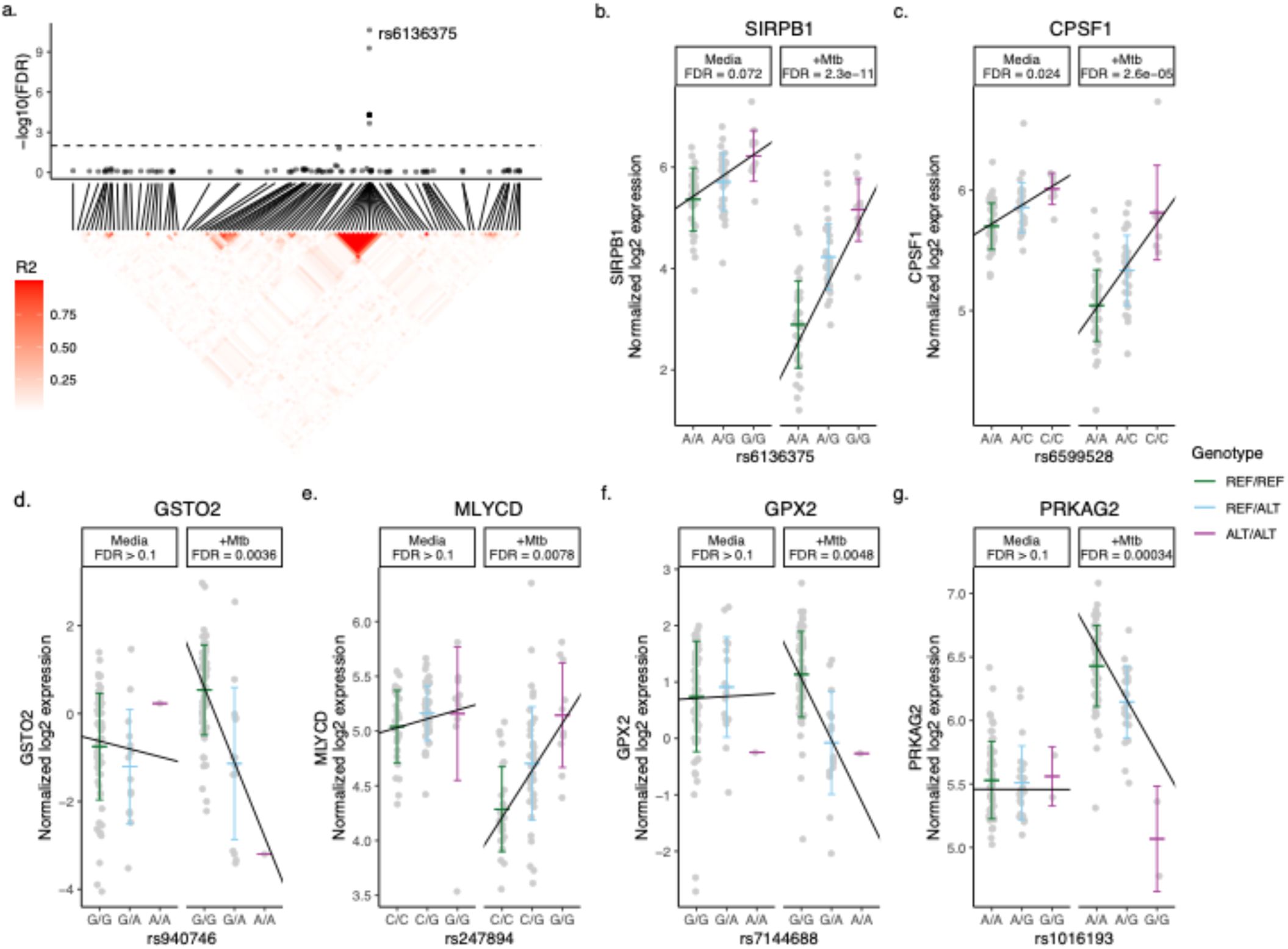
Mtb-dependent eQTL Monocytes Expression Plots. eQTLs were defined by significance in the Mtb condition (FDR < 0.01) and interaction term (FDR < 0.1) models but not in media. (a, top) eQTL significance for SNPs within 1 Mb of SIRPB1. X-axis indicates position in chromosome 20. Horizontal dashed line indicates FDR = 0.1 in the interaction model and the lead SNP is labeled. (a, bottom) Heatmap indicating R^2^ linkage disequilibrium (LD) for SNPs in this region. (b-g) eQTL plots illustrate the relationship between genotype (x-axis) and log2 expression (y-axis) of the indicated genes. Each line indicates a linear fit derived from an additive regression model that includes an interaction term (Mtb_infection:Genotype), adjusting for age and sex. False discovery rates (FDR) for each eQTL are reported for the Mtb-uninfected media condition and the Mtb-infected condition, not the interaction model, and are presented to illustrate the effects of Mtb stimulation and host genotype. The lead eQTLs (b) rs6136375 for SIRPB1 and (c) rs6599528 for CPSF1 may have minor genotype effects independent of Mtb stimulation. (d) rs940746 for GSTO2 and (e) rs247894 for MLYCD highlight loci where no genetic effects were observed in the media condition (FDR > 0.9), but which become significant upon Mtb infection. (f) rs7144688 for GPX2 and (g) rs1016193 for PRKAG2 highlight loci where the directionality of genotype effects differed between the media and Mtb conditions. ALT = alternative allele, eQTL = expression quantitative trait loci, FDR = false discovery rate, Mb = megabase, Mtb = *Mycobacterium tuberculosis,* REF = reference allele, SNP = single nucleotide polymorphism

**Table 2.** Mtb-dependent eQTL genes and 16 lead eQTLs.

| Gene | eQTL SNP ID | SNP position* | Genotype | MAF | t-statistic | FDR** |
| --- | --- | --- | --- | --- | --- | --- |
| **SIRPB1** | rs6136375 | Chr20:1896100 | A>G | 0.383 | 5.146 | 0.001 |
| **CHMP1B** | rs7241199 | Chr18:11857066 | G>A | 0.344 | -4.414 | 0.007 |
| **MRPL55** | rs74597096 | Chr1:228317858 | A>C | 0.100 | -4.388 | 0.007 |
| **TIMM44** | rs12610448 | Chr19:8532219 | A>G | 0.172 | -4.335 | 0.007 |
| **PRKAG2** | rs1016193 | Chr7:150412100 | A>G | 0.188 | -4.193 | 0.01 |
| **NDUFAF4** | rs78956122 | Chr6:96605485 | G>A | 0.066 | -3.833 | 0.031 |
| **GPX2** | rs7144688 | Chr14:64613473 | G>A | 0.107 | 3.607 | 0.035 |
| **FANCA** | rs550516418 | Chr16:89626136 | A>T | 0.051 | -3.56 | 0.037 |
| **TRMT13** | rs7555025 | Chr1:100997680 | A>G | 0.066 | -3.47 | 0.037 |
| **MLYCD** | rs247894 | Chr16:84479189 | C>G | 0.418 | -3.469 | 0.037 |
| **GSTO2** | rs940746 | Chr10:106763758 | G>A | 0.090 | 3.477 | 0.049 |
| **KCNK5** | rs2815125 | Chr6:39194533 | A>G | 0.164 | -3.369 | 0.059 |
| **BMP6** | rs55966428 | Chr6:8829127 | A>G | 0.148 | 3.302 | 0.059 |
| **CPSF1** | rs6599528 | Chr8:145603114 | A>C | 0.238 | -3.29 | 0.063 |
| **PIBF1** | rs7994794 | Chr13:73086549 | A>G | 0.328 | 3.251 | 0.067 |
| **MAS1** | rs6924573 | Chr6:160286188 | C>A | 0.279 | 3.222 | 0.069 |

An additive linear regression model, including an interaction term (Mtb\_infection:Genotype), is employed to calculate the t-statistics of eQTLs. The selection of Mtb-dependent eQTLs is based on FDR values to account for false positives resulting from multiple testing. eQTLs with an FDR value below 0.1 are considered statistically significant Mtb-dependent eQTLs, and only the lead eQTLs are listed in this table.
FDR = false discovery rate, MAF = minor allele frequency, p = p-value, SNP = single nucleotide polymorphism.
\* GRCh37
\*\* The FDR values of the interaction model

### Network and enrichment analyses implicate Mtb-dependent eQTL genes in fatty acid and glutathione metabolism, along with other cellular metabolic processes

To investigate the functional roles of 16 Mtb-dependent eQTL genes, we constructed a network of their protein-level interactions using the STRING database [17] and identified two high-confidence edges (STRING score > 700, Figure 4). To gain further insight into the functional roles of these genes, we performed hypergeometric mean enrichment pathway analysis of the 16 Mtb-dependent eQTL genes using the Human Molecular Signatures Database (MsigDB v2023.1, Table S3) [18]. In the C2 canonical pathway, we observed significant enrichment of gene sets related to the fatty acid and glutathione metabolism for *GPX2, GSTO2, MLYCD,* and *PRKAG2* (FDR < 0.1). Additionally, within the C5 gene ontology biological pathways, we found associations of *BMP6, CHMP1B, FANCA, MAS1, MLYCD, PRKAG2,* and *PIBF1* with 52 gene sets related to fatty acid, lipid, and organic hydroxy metabolism as well as mitotic spindle (Table S3, FDR < 0.1). In summary, our network and enrichment analyses of the 16 Mtb-dependent eQTL genes revealed their involvement in multiple pathways associated with fatty acid metabolism, glutathione metabolism, and other cellular metabolic processes.

**Figure 4.**
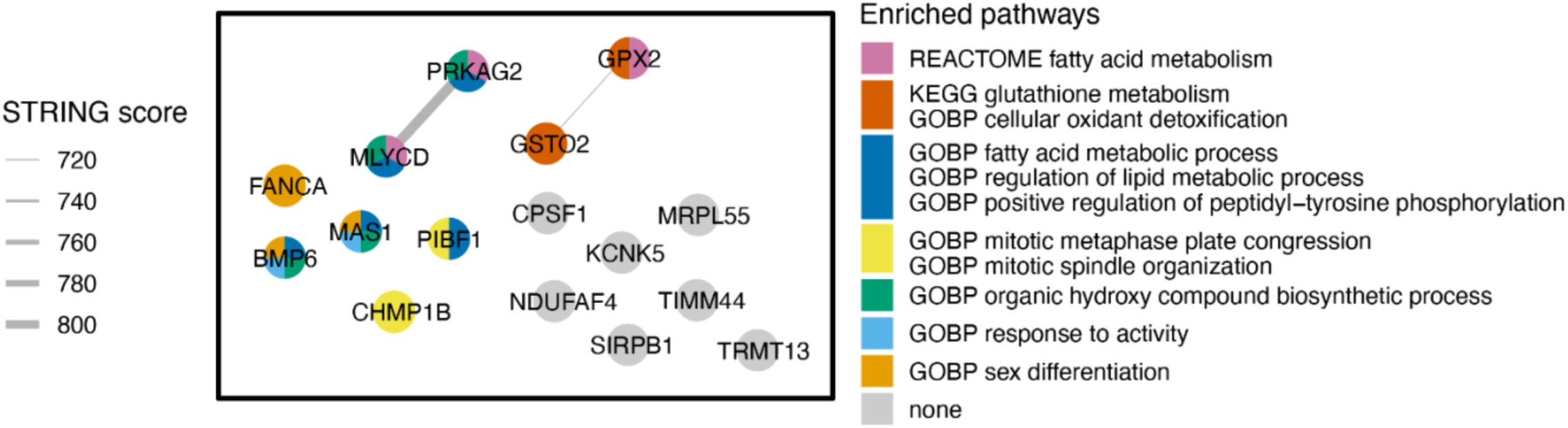
Network and Gene Enrichment Pathway Analysis. The left-side box in the figure illustrates the STRING network of protein-protein interactions among 16 Mtb-dependent eQTL genes. Each node denotes a Mtb-dependent eQTL gene and each line indicates a protein-level interaction. Nodes are colored by hypergeometric mean enrichment pathway analysis with MSigDB canonical pathways and gene ontology. PRKAG2 and MLYCD are functionally related in regulation of fatty acid oxidation and metabolic process. GPX2 and GSTO2 are functionally related in glutathione metabolism. GOBP = Gene Ontology Biological Process, KEGG = Kyoto Encyclopedia of Genes and Genomes, STRING = Search Tool for the Retrieval of Interacting Genes/Proteins.

### Polymorphisms in Mtb-dependent eQTL genes and association with resistance to TST/IGRA conversion in TB household contacts

We examined the clinical significance of Mtb-dependent eQTL genes and variants by testing their association with resistance to TST/IGRA conversion in highly exposed HHCs of pulmonary TB subjects. We conducted a candidate gene association study with a previously collected extended sample size (RSTR [case], n=74) versus (LTBI [control], n=189) for each Mtb-dependent eQTL gene, adjusting for age, sex, and kinship using a generalized linear mixed model (GENESIS) [11]. One of the Mtb-dependent eQTLs, rs6599528 (associated with the cis-expression of *CPSF1*), was significantly associated with the RSTR phenotype where the adjusted odds of being RSTR increased by 1.63 times for each copy of the minor allele at this locus (p= 0.04, Table 3). We also examined whether SNPs within the up- and down-stream 5-kilobase (kb) flanking sequences of each of the 16 Mtb-dependent eQTL genes were associated with the RSTR outcome. We identified 26 SNPs in 7 genes associated with clinical RSTR status (p < 0.05; Table S4), the majority of which were found in *PRKAG2* (n= 20), with 6 having previously been reported [13, 19]. None of these findings remained significant after correction for multiple comparisons. Taken together, our results suggest that Mtb-dependent eQTLs and SNPs in their gene region may be associated with TB clinical outcomes.

**Table 3.** Association of CPSF1 with clinical resistance to TST/IGRA conversion.

| SNP ID | SNP position | Genotype | Genotype Frequency |  | OR* | *p* | R<sup>2</sup>** |
| --- | --- | --- | --- | --- | --- | --- | --- |
|  |  |  | RSTR | LTBI |  |  |  |
| Mtb-dependent eQTL |  |  |  |  |  |  |  |
| rs6599528 | 8:145603114 | A>C | A/C: 26/74<br>C/C: 7/74 | A/C: 57/189<br>C/C: 7/189 | 1.63 | 0.048 | 1.0 |
| SNPs within ± 5kb flanking sequences of CPSF1 |  |  |  |  |  |  |  |
| rs147468511 | 8: 145625831 | G>A | G/A: 15/74<br>A/A: 0/74 | G/A: 19/189<br>A/A: 0/189 | 2.42 | 0.042 | 0.028 |
| rs4925820 | 8:145637148 | C>A | C/A: 38/74<br>A/A: 18/74 | C/A: 94/189<br>A/A: 34/189 | 1.33 | 0.181 | 0.221 |
| rs736676 | 8:145639320 | C>G | C/G: 28/74<br>G/G: 5/74 | C/G: 67/189<br>G/G: 16/189 | 1.00 | 0.991 | 0.241 |
| rs75920625 | 8:145639654 | A>G | A/G: 32/74<br>G/G: 1/74 | A/G: 68/189<br>G/G: 17/189 | 0.82 | 0.419 | 0.008 |
| rs1871534 | 8:145639681 | C>G | C/G: 17/73<br>G/G: 1/73 | C/G: 30/187<br>G/G: 3/187 | 1.56 | 0.206 | 0.056 |
| rs2272662 | 8:145639726 | A>G | A/G: 10/70<br>G/G: 0/70 | A/G: 24/183<br>G/G: 0/183 | 1.09 | 0.841 | 0.097 |

\* In an additive model, the odds ratio (OR) measures the change in the odds of being RSTR with each additional copy of the minor allele.
\*\* The R<sup>2</sup> value of linkage disequilibrium (LD) from Mtb-dependent eQTL to the designated SNP within the Mtb-dependent eQTL gene
IGRA = interferon-γ release assay, kb = kilobase, LTBI = latent tuberculosis infection, OR = odds ratio, *p* = *p*-value, RSTR = resister or resistant to TST/IGRA conversion after high TB exposure, SNP = single nucleotide polymorphism, TST = tuberculin skin test.

### Mtb-dependent eQTLs associate with monocyte cytokine expression

Under inflammatory conditions, genetic polymorphisms of pro-inflammatory cytokines, such as tumor necrosis factor (TNF), interleukin (IL)-1β, IL-6, or type I interferons (IFN-β), have been associated with susceptibility to TB in humans [20–23]. We examined whether Mtb-dependent eQTLs are associated with the expression levels of these cytokines. We identified 8 of the 29 Mtb-dependent eQTLs that were associated with Mtb-induced cytokine expression in monocytes (additive regression model adjusted for age and sex, p < 0.05, Figure 2a). Among these, 4 eQTLs (rs1016193 [*PRKAG2*], rs12610448 [*TIMM44*], rs55966428 [*BMP6*], rs7241199 [*CHMP1B*]) were associated with altered *IFNB1* gene expression in response to Mtb infection (p < 0.05, Figure 5a-c and Figure 6c). The remaining 4 eQTLs (rs247894 [*MLYCD*], rs7994794 [*PIBF1*], rs6599528 [*CPSF1*], rs6924573 [*MAS1*]) were significantly associated with altered *TNF*, *IL6* and/or *IL1B* expression levels between Mtb-infected and uninfected monocytes (p < 0.05, Figure 5d-h, Table S5). We also observed that 6 of the 8 eQTL genes additionally had variants that were associated with clinical RSTR outcomes (Figure 2a). These findings indicate that Mtb-dependent eQTL genes identify host genes that may regulate Mtb-induced cytokine induction *in trans* and potentially contribute to clinical resistance.

**Figure 5.**
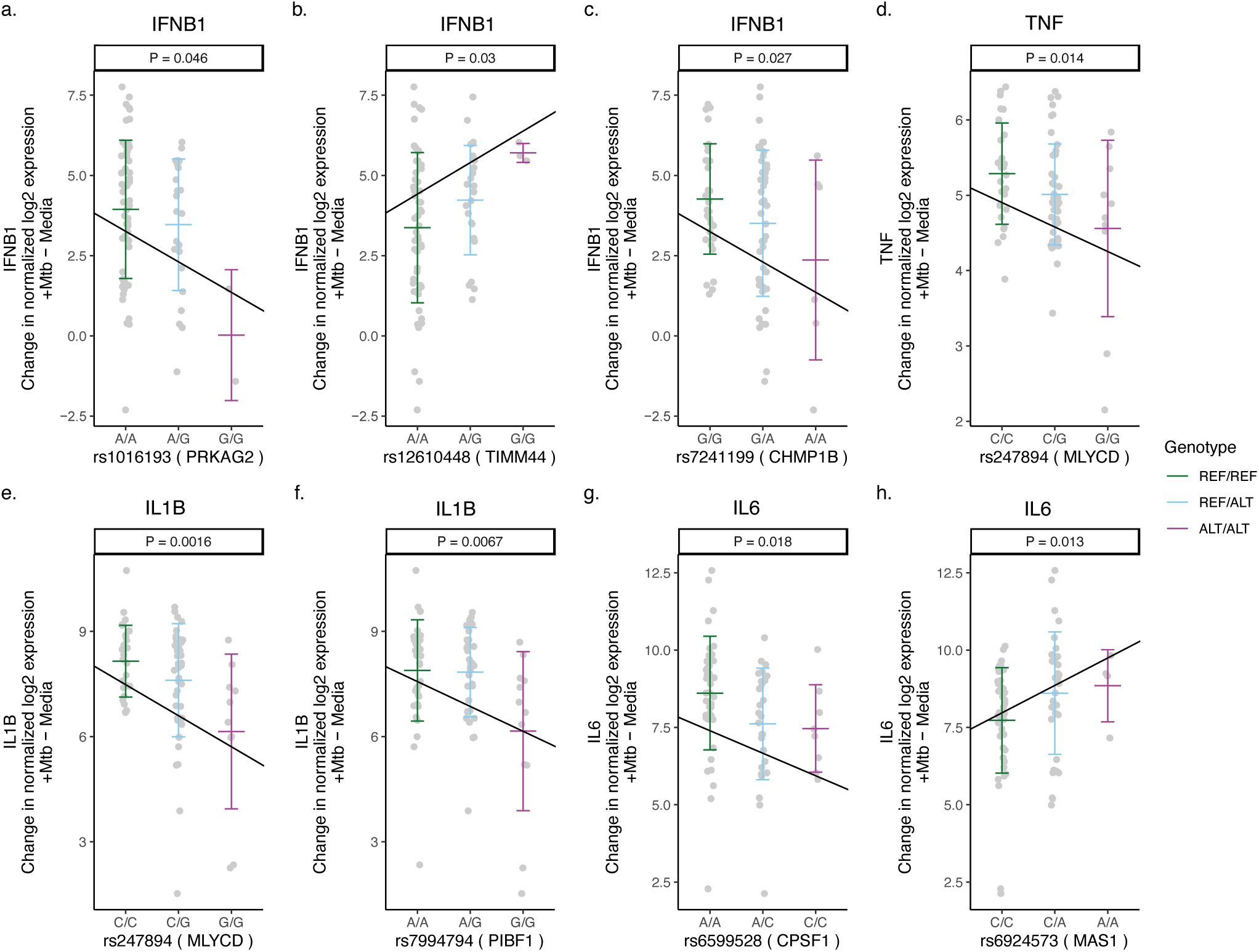
Mtb-dependent eQTLs associated with Mtb-induced cytokine expression in monocytes. Three Mtb-dependent eQTLs (a-c), rs1016193, rs12610448, and rs7241199, were associated with the expression of PRKAG2, TIMM44, and CHMP1B *in cis*, respectively, and with the regulation of IFNB1 upon Mtb infection. (d) rs247894 was associated with the cis-regulation of MLYCD and the regulation of TNF expression. (e) rs247894 and (f) rs7994794 were associated with the cis-regulation of MLYCD and PIBF1 expression, respectively, as well as with the regulation of IL1B. (g) rs6599528 and (h) rs6924573 were associated with the cis-regulation of CPSF1 and MAS1 expression, respectively, along with the regulation of IL6. The slope of the lines indicates the ratio of the effect estimate to the standard error derived from a linear regression model, which estimates the relationship between the number of minor alleles and the differences in cytokine expression (normalized log2 [Mtb-infected – uninfected relative cytokine expression]), after accounting for age and sex (p < 0.05). ALT = alternative allele, REF = reference allele

**Figure 6.**
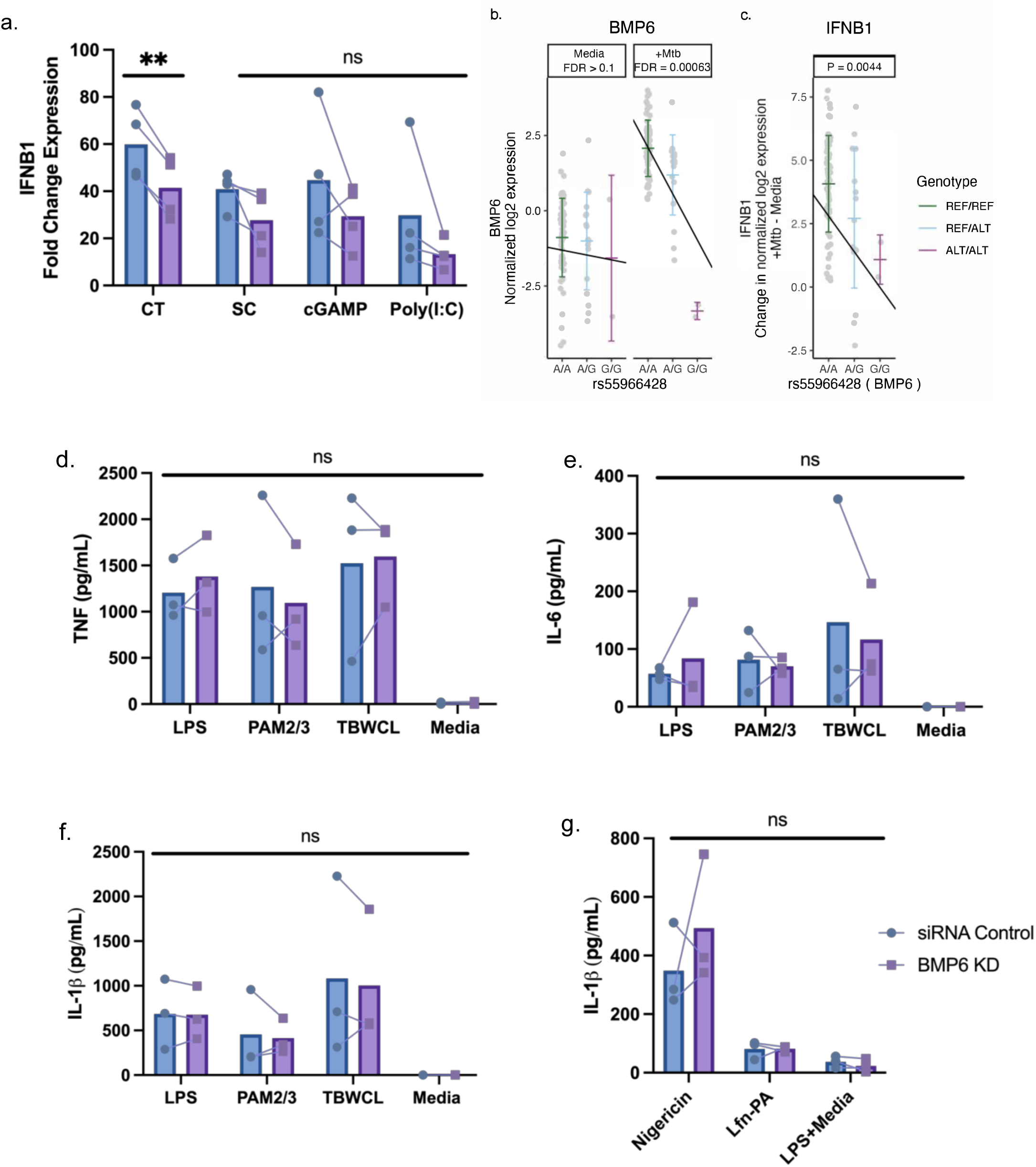
BMP6 expression is associated with host genotype in Mtb-infected cells and is required for maximal IFNB1 responses in monocytes following DNA-ligand stimulation. (a) The fold change expression of IFNB1 was significantly reduced in BMP6-silenced THP1 cells after 4h of stimulation with 4 μg/mL calf thymus DNA ligand, compared to siRNA control cells. BMP6-silenced cells stimulated with 1 μg/mL of super coiled plasmid DNA, 20 μM of cGAMP, or 10 μg/mL of poly(I:C) exhibited statistically non-significant decreases in IFNB1 induction compared to siRNA control cells. The fold change expression of IFNB1 was measured by qPCR and normalized against the IFNB1 expression from lipofectamine without DNA/RNA ligand to control background IFNB1 induction from lipofectamine. For TLR-specific stimulation, BMP6-silenced and siRNA control THP-1 cells were stimulated with LPS, PAM2/PAM3, Mtb whole cell lysate, or media for 24h. (b) In the Ugandan HCC cohort, the minor allele (G) of rs55966428 was associated with decreased expression of BMP6 following 6h of Mtb infection in primary monocyte data from HHCs. Host genotype (x-axis) and normalized log2 expression (y-axis) of BMP6 for the rs55966428 Mtb-dependent eQTL with false discovery rate (FDR) reported separately for the media and Mtb-stimulated conditions. (c) In the same cohort, the minor allele (G) of rs55966428 was associated with decreased IFNB1 expression following 6h of Mtb infection in primary monocyte data. This plot shows genotype on the x-axis and differences in IFNB1 expression on the y-axis (normalized log2 [Mtb-infected – uninfected relative IFNB1 expression]), adjusting for age and sex. (d) TNF, (e) IL-6, and (f) IL-1β supernatant levels were measured using ELISA. To assess (g) inflammasome-mediated IL-1β response, nucleofected cells were primed with LPS for 2h and treated with nigericin for NLRP3-specific stimulation, *Burkholderia thailandensis* needle protein for 4h for NLRC4-specific stimulation. Needle protein was administered at 8 ng/ml in conjunction with 16 ng/ml *Bacillus anthracis* protective antigen. The IL-1β supernatant levels were measured by ELISA. cGAMP = cyclic guanosine monophosphate-adenosine monophosphate, CT = calf thymus DNA, LPS = lipopolysaccharide, media = media control (RPMI + 10% FBS), PAM2 = Pam2CSK4, PAM3 = Pam3CSK, poly(I:C) = polyinosinic acid-polycytidylic acid, SC = super coiled plasmid DNA, TBWCL = Mtb whole cell lysates, TLR = toll-like receptor. **p < 0.01 tested by paired t-test, ns: not significant.

### *BMP6* expression is associated with Mtb and DNA-ligand induced IFNB expression in monocytes

To further test our hypothesis that Mtb-dependent eQTL genes regulate cytokine pathways in monocytes, we used gene silencing methods to investigate potential mechanisms. Among 16 Mtb-dependent eQTL genes, we prioritized genes based on three criteria: (i) more pronounced slope changes between Mtb-infected and uninfected conditions, (ii) significant associations with cytokine gene expression, and (iii) significant associations with the clinical RSTR outcomes. From this analysis, *BMP6, CHMP1B, KCNK5, PIBF1, PRKAG2,* and *TIMM44* were initially selected for gene silencing experiments. After assessing siRNA knockdown efficiency, we ultimately focused on *BMP6, PRKAG2,* and *TIMM44* to investigate their role in cytokine signaling experiments.

After silencing expression of *BMP6, PRKAG2,* and *TIMM44*, we stimulated PMA-differentiated THP-1 cells with DNA/RNA, toll-like receptor (TLR), or inflammasome ligands. We observed a decrease in *IFNB1* expression after 4h of stimulation with 4 μg/mL sheared calf thymus DNA in *BMP6*-knockdown cells compared to the control group (Figure 6a). This expression pattern aligns with the HHC primary monocyte data, where the minor allele (G) of rs55966428 was associated with a significant reduction in the expression of both *BMP6* (Figure 6b) and *IFNB1* (Figure 6c) after 6h of Mtb infection. Although the other DNA/RNA ligands showed a similar trend of lower *IFNB1* expression, the results were not statistically significant. In contrast, we did not find significant differences in *IFNB* expression for *PRKAG2* or *TIMM44* (Figure S3a). There were no significant differences in TNF, IL-6, or IL-1β secretion observed between the *BMP6*, *PRKAG2*, and *TIMM44*-silenced cells and the siRNA controls (Figure 6d-g, Figure S3b-e). Collectively, our data suggests that *BMP6* regulates the expression of *IFNB1* following Mtb and DNA ligand stimulation, but not TLR- or inflammasome-induced cytokine expression.

## DISCUSSION

Myeloid cells play a critical role to restrict Mtb replication through intrinsic microbicidal pathways and by stimulating downstream cellular responses, but the genetic factors that determine Mtb infection and disease outcomes remains largely unknown. While we previously identified 260 differentially expressed genes in monocytes in response to Mtb infection and clinical TB phenotypes [14], this analysis was limited in fully capturing the genetically encoded monocyte responses following Mtb infection. In the current study, using genome-wide eQTL mapping in Mtb-infected monocytes, we identified 29 Mtb-dependent eQTLs in 16 genes. A subset of these eQTLs is also associated with Mtb-induced cytokine gene expression and/or clinical resistance to TST/IGRA conversion (RSTR phenotype). The expression profiles of these Mtb-dependent eQTL genes and their associated pathways indicate the involvement of diverse inflammatory and cellular metabolic processes, potentially influencing the mechanisms underlying resistance or susceptibility to Mtb infection.

Our primary findings suggest that *BMP6* (bone morphogenetic protein 6), a member of the transforming growth factor (TGF)-β superfamily, is involved in the genetic regulation of IFN-β and may influence antimicrobial responses. *BMP6* plays a critical role in tissue remodeling and organogenesis [24, 25]. The TGF-β and BMP pathways have an antagonistic relationship due to shared receptor structures and signaling mechanisms [26, 27], which have been observed in conditions like pulmonary fibrosis [28, 29] and malignancies [30–32]. In myeloid cells, *BMP6* distinguishes itself with its unique pro-inflammatory function, enhancing antimicrobial activities, while other BMPs (*BMP2*, *BMP4*, and *BMP7*) contribute to anti-inflammatory functions and the promotion of the M2 macrophage phenotype [33]. In murine models, BMP6 treatment activates NF-kB signaling, induces IL-1β and TNF expression, and increases the expression of inducible nitric oxide synthase (iNOS), resulting in elevated nitric oxide production and enhanced microbicidal effects [34, 35]. In contrast, human data has shown that BMP6 induces the expression of RIG-I-like receptors (RLRs) like RIG-I and MDA5, which regulate IFN-β and restrict Zika virus replication [36]. We found that individuals with the minor allele (G) at rs55966428 showed decreased expression of both *BMP6* and *IFNB1*. Additionally, *BMP6* silencing in THP-1 cells resulted in a significant decrease in IFNB1 expression upon DNA ligand stimulation, partially validating *BMP6’s* role in regulating IFN-β induction via DNA sensing pathways. In the context of Mtb infection, *in vivo* data from animal infection models reveal that activation of cytosolic cGAS- or RLR-mediated sensing pathways triggers a robust type I IFN response that impairs host resistance to Mtb infection [37–39]. Conversely, activation of other cytosolic pathways during Mtb infection, such as AIM2, NOD2, and NLRP3, promotes the production of protective inflammatory cytokines [40]. Therefore, further investigation is warranted to explore the precise role of *BMP6* in either stimulating or inhibiting host-protective pro-inflammatory functions, as well as the regulatory factors involved in DNA/RNA sensor-dependent IFN-β responses, along with their impact on antimicrobial properties and clinical resistance to Mtb infection.

We also found evidence suggesting Mtb may regulate monocyte gene expression through mRNA processing. *CPSF1* (cleavage and polyadenylation specific factor 1) is crucial for mRNA maturation and alternative 3’ untranslated region isoform generation [41]. The involvement of *CPSF1* in various human diseases, including cancers [42–44] and ocular disorders [45, 46] has been studied, but its role in TB susceptibility remains unexplored. Importantly, our genetic analysis revealed that the minor allele (C) at rs6599528 was associated with a 1.63-fold increase in the odds of being RSTR, potentially indicating a protective property of the C allele against TST/IGRA conversion. Upon Mtb infection, the majority of individuals who carry the major allele (A) has decreased expression of *CPSF1*; while no changes in its expression among individuals with recessive genotype (C/C). These findings suggest that the genotypic regulation of *CPSF1* via rs6599528 may potentially mediate alternative splicing of host transcripts that influence susceptibility to Mtb infection. Among potential host pathways that mediate this protection, recent attention has focused on the alternative role of *CPSF1* as an E3 ubiquitin ligase that degrades the transcription factor hypoxia-inducible factor (HIF)-1α. HIF-1α is essential for the IFN-γ-driven macrophage immunometabolic response [47, 48] and its activation results in increased *IL6* expression [49]. Our findings indicate that individuals with the homozygous recessive genotype (C/C) at this locus, which is linked to increased *CPSF1* expression, are predominantly observed in the RSTR group, which is characterized by a lack of response to IFN-γ activation signals (e.g., IGRA positivity). Increased *CPSF1* expression among RSTRs, which is predicted to result in HIF-1a degradation, is consistent with IFN-γ-independent protective mechanisms [50, 51]. Interestingly, in our data, *IL6* expression is elevated in resting monocytes carrying the minor allele (Figure S4h). However, the genetic effects of rs6599528 on *IL6* expression become non-significant following Mtb infection, indicating that IL-6 regulation is influenced by more complex and multifaceted factors. Therefore, further investigations are needed to elucidate the complex influence of rs6599528 on pro-inflammatory signaling, antimicrobial mechanisms against Mtb, and clinical resistance outcomes.

Mtb has evolved strategies to manipulate macrophage metabolism by utilizing host fatty acids and cholesterol as its primary energy sources [52–54]. We identified 6 Mtb-dependent eQTL genes, including *PRKAG2*, *MLYCD*, *BMP6*, *MAS1*, *PIBF1*, and *GPX2*, that are involved in fatty acid metabolism. The heterotrimeric AMP-activated protein kinase (AMPK) contains one kinase (AMPK-α) and two regulatory domains (AMPK-β and AMPK-γ), which upon activation promotes catabolic processes such as fatty acid oxidation (FAO) [55, 56] by phosphorylation of central metabolic enzymes [57]. Genetic variants in *PRKAG2*, which encodes one AMPK-γ isoform, are associated with the RSTR phenotype [13]. Moreover, we identified a conserved enrichment of gene sets associated with carbon metabolism and the transcriptional response to free fatty acids (FFAs) across independent RSTR cohorts in Uganda and South Africa [13]. Although the *PRKAG2* Mtb-dependent eQTLs identified here are distinct from *PRKAG2* polymorphisms that are associated with the RSTR phenotype, our findings further support a role for *PRKAG2* and AMPK in measurable *in vitro* and clinical TB outcomes that is under selective pressure. To further support host lipid metabolism during monocyte Mtb responses, the *MLYCD* Mtb-dependent eQTL indicates host-genotype dependent responses to Mtb that may impact lipid synthesis. Malonyl-CoA decarboxylase (MCD), encoded by *MLYCD*, catalyzes the decarboxylation of malonyl-CoA [58–60]. Since malonyl-CoA is both an intermediate during lipid synthesis and an allosteric inhibitor of carnitine palmitoyltransferase 1 (CPT1)-mediated FAO, MCD activity results in the inhibition of lipid synthesis and the promotion of FAO. Although the direct impact of *MLYCD* downregulation in response to Mtb infection has not been studied, it is likely to disrupt FAO. Consistent with the predicted effect that *MLYCD* downregulation (and thus malonyl-CoA accumulation) may have towards Mtb virulence, Mehrotra et al. [61] measured increased malonyl-CoA levels following Mtb infection that was specific to virulent strains and resulted in net fatty acid synthesis. Further inhibition of FAO induces HIF-1α and excessive production of reactive oxygen species (ROS) from the mitochondria [62]. As a result, the excessive HIF-1α and ROS promote the transcription of glycolytic enzymes and inflammatory mediators, including IL-1β, IL-6 and TNF, facilitating additional antimicrobial responses [63, 64]. Our data demonstrate that monocytes with homozygous dominance at rs247894 exhibit reduced *MLYCD* expression *in cis* but increased *IL1B* and *TNF* expression *in trans* in response to Mtb infection. These findings suggest that the downregulation of *MLYCD* contributes to the host protective pro-inflammatory properties. However, reduced FAO also leads to the accumulation of lipids that can serve as a nutrient source for Mtb growth [65, 66]. The conflicting associations observed in *MLYCD* suggest an ongoing evolutionary battle within host cells, necessitating further investigation into its role in immunometabolic function and host-protective response. Paradoxically, macrophages also employ redox regulation mechanisms, primarily reliant on glutathione metabolism, to prevent oxidative damage caused primarily by NADPH-oxidase-driven hydrogen peroxide [67]. Our findings support the involvement of glutathione-mediated antioxidant enzymes, *GPX2* (glutathione peroxidase 2) [68–70] and *GSTO2* (glutathione S-transferase omega 2) [71, 72], in redox regulation during Mtb infection. These enzymes, through genetically regulated expression that is induced by Mtb, may influence antioxidant defense or bacterial clearance [73, 74]. Together, our study highlights the complex interplay between immunometabolic pathways, genetic factors, and Mtb infection outcomes, emphasizing the necessity for further research to elucidate the roles of Mtb-dependent eQTL genes and their impact on immunometabolic function and host protection in the context of Mtb infection.

Prior studies of Mtb-dependent eQTLs illuminate potential areas of convergence of findings. Previously, Barreiro et al. [75] identified 102 eQTLs in Mtb-uninfected and 96 in Mtb-infected monocyte-derived dendritic cells (DCs) using a similar study design with some significant differences, including cell type (DCs versus monocytes), age (adults versus range of ages), sex (male versus both male and female), population background (White versus Ugandan), time point (18h versus 6h), and transcriptomic method (expression array versus RNAseq). In addition, different statistical criteria were used to define “response eQTLs” (FDR significance in one condition but not the other versus Mtb-infection:genotype interaction model) and distance of cis-regulatory effects (200kb proximity from the TSS, versus 1Mb). Despite no identical eQTLs or eQTL genes being found, both studies found several genes within the same family. For example, *BMP1*, a response eQTL gene in uninfected DCs, shared the same gene family with our Mtb-dependent eQTL gene, *BMP6*. Unlike the rest of the BMP family, BMP1 does not belong to the TGF-β superfamily; instead, it acts as a procollagen C-proteinase in collagen maturation and indirectly contributes to TGF-β upregulation by activating matrix metalloproteinase 2 (MMP2) and MMP9, influencing tissue remodeling [76, 77]. In addition, Mtb-infected DCs exhibited *CPSF3* and *NDUFAF2* as response eQTL genes, belonging to the same gene family as our Mtb-dependent eQTL genes, *CPSF1*, and *NDUFAF4*, respectively. Little research has focused on *CPSF3*, but like *CPSF1*, it is thought to be involved in mRNA processing and may contribute to the regulation of gene expression in response to diverse pathogenic microorganisms, such as *Toxoplasma gondii* [78], *Plasmodium falciparum* [79], and *Cryptosporidium* [80] infection. Finally, *NDUFAF4* plays an essential role in the assembly and maturation of the NADH:ubiquinone oxidoreductase complex (complex I) in the mitochondrial respiratory chain [81, 82], linked with the late-stage assembly factor *NDUFAF2* [83, 84], crucial for energy production through oxidative phosphorylation [85]. The convergence of these findings adds weight to the results and encourages further exploration of shared genes or pathways.

This study has several limitations. First, the ‘Mtb-dependent eQTLs’ identified may not be specific to Mtb infection since our stimulation condition did not include other pathogens or ligands, an area for future investigation. Additionally, our findings using peripheral blood monocytes may not extend to other tissues and cell types at the site of Mtb pathogenesis such as alveolar macrophages where variants or Mtb-dependent eQTL genes may be distinct. Finally, although our siRNA silencing experiments and cytokine gene association analysis suggest a plausible causal effect that Mtb-dependent eQTL gene expression has on cytokine responses following Mtb-infection, future advanced functional assays will explore the regulatory roles of eQTLs’ promoter/enhancer elements that mediate these responses.

In summary, this study demonstrates the robust application of genome-wide eQTL mapping to investigate genetic regulation of the host response against Mtb. Our findings highlight significant associations between Mtb-dependent eQTL genes and variants and alterations in the immune response to Mtb infection, as well as potential regulatory interactions between specific eQTLs and Mtb-induced cytokines. These insights into immunogenetic mechanisms provide valuable targets for host-directed therapies and TB control.

## METHODS

### Overview of study design and sampling

Between 2002 and 2012 (phase 1), we enrolled 872 culture-confirmed pulmonary TB index cases and their 2,585 HHCs in the Kawempe Community Health Study in Kampala, Uganda [86]. HHCs were defined as individuals who had lived in the same household as the TB index case for at least seven consecutive days in the preceding 3 months. At baseline and every 3-6 months thereafter for up to 24 months, all HHCs were evaluated for evidence of LTBI using TST. Between 2014 and 2017 (phase 2), a subgroup of these HHCs were re-contacted after 8-10 years of follow-up, and serial TST and IGRA were completed for TB screening when PBMCs were also collected. Detailed recruitment and screening procedures for this Ugandan HHC cohort have been previously described [11, 15, 86].

### CD14+ Monocyte isolation, Mtb infection, and RNA sequencing

Cryopreserved PBMCs (n=115) were thawed and resuspended in RPMI 1640 medium (Gibco) supplemented with 10% heat-inactivated fetal bovine serum (FBS) and 50 ng/mL recombinant human monocyte colony-stimulated factor (M-CSF) for 24h. CD14+ monocytes were enriched from PBMCs by magnetic separation (Monocyte Isolation Kit II, Miltenyi Biotec) and incubated in RPMI-10 with M-CSF for 24h. Monocytes were then infected with H37Rv Mtb at a multiplicity of infection (MOI) of 1 or media alone and incubated at 37 °C with 5% CO2 for 6h. After incubation, both the media-only and Mtb-infected monocytes were lysed in TRIzol (Invitrogen), and RNA was isolated using the RNeasy Mini Kit according to the manufacturers’ instructions (Qiagen). The RNA samples were quantified using Nanodrop 8000 instrument (Thermo Scientific), and their quality was measured by TapeStation (Agilent) (RNA Integrity Number ≥ 8.0). Next, cDNA libraries were prepared with random hexanucleotide primers and rRNA depletion using SMARTer RNAseq Kit (Takara), followed by sequencing on Illumina HiSeq 2500 and Novaseq 6000 platforms as previously described [14].

### Sequence data processing

Sequences were processed as described previously [14]. Briefly, sequences were aligned to the GRCh38 reference genome using STAR 2.6.0 [87] and quantified read counts using RSEM 1.3.0 [88]. We excluded 4,691 genes (Figure 1) that were non-protein coding, not reliably annotated, or located on a sex chromosome. The latter would have complicated assessment of deviation from Hardy-Weinberg equilibrium (HWE) due to males being hemizygous. This left 13,439 genes, which we then trimmed-mean of M-values (TMM) normalized and converted to log2 counts per million (CPM) using limma [89]. In parallel, genome-wide genotyping of 263 HHCs was performed using the Illumina MEGA^EX^ array. We successfully genotyped 1,002,430 SNPs, of which we filtered out 731,662 SNPs that deviated from HWE (P < 10^-6^) and had a minor allele frequency (MAF) of less than 0.05.

### eQTL analysis

To identify genetic variants that regulate mRNA expression in a cis-acting manner, we tested for associations between transcript expression levels and genotypes at SNPs located within a 1Mb window centered on the genes’ transcription start sites (TSS). As we lacked substantial power for trans analysis due to the small sample size, we focused solely on cis-eQTLs. Initially, we performed an additive linear regression of genotypes on log-transformed gene expression level, adjusted for age and sex in either Mtb-infected or uninfected monocytes. The R package MatrixEQTL was used to perform all regressions [90]. We estimated FDR by using a Fisher’s exact test to correct for multiple testing by the Benjamini–Hochberg method to control for false positives. Significant eQTLs were defined as those with an FDR < 0.01 in Mtb-infected or uninfected monocytes.

For the 1,567 eQTLs identified in Mtb-infected monocytes, we further introduced an interaction term (Mtb-infection:genotype) in the main additive regression model to uncover whether the genetic effect of eQTLs on the level of gene expression differed by Mtb infection status. We applied a loose cut-off of FDR < 0.1 in the interaction model, given the small number of eQTLs included in this analysis [91]. From the 32 eQTLs that showed significance in Mtb-infected condition and the interaction model, we removed 3 eQTLs that overlapped with those identified in uninfected condition to capture only those that regulate target genes in response to Mtb infection, which we named “Mtb-dependent eQTLs”. Finally, we defined the lead eQTLs per gene as the SNP that was most significantly associated (with the lowest FDR value) with the expression of that gene (Figure 2).

### Mapping cytokine expression levels

The associations between genotype and cytokine expression were assessed for each of the 16 lead Mtb-dependent eQTLs. Specifically, we examined the expression levels of *TNF, IL6, IL1B*, and *IFNB1* in Mtb-infected and uninfected monocytes, as genetic variation in these cytokine genes has previously been linked to TB susceptibility [20–23]. We used a linear regression model to regress the number of minor alleles on the differences in cytokine expression between Mtb-infected and uninfected monocytes (normalized log2 [Mtb-infected – uninfected relative expression]), while adjusting for age and sex, at a significance level of 0.05.

### Mtb-dependent eQTL gene association study

We conducted association analyses to determine whether a particular eQTL genotype co-occurs with a RSTR clinical phenotype more often than would be expected by chance. We calculated odds ratios (ORs), adjusted for age and sex as well as kinship using a quasi-likelihood approximation to the generalized linear mixed model (GLMM) (GENESIS, R package) [92]. Statistical comparison of demographic and epidemiologic data between RSTR and LTBI groups were conducted at a 2-sided significance level of 0.05 using Stata/IC 15.1 (StataCorp LLC) [93].

### Integrative pathway enrichment analysis

To examine potential protein-level interactions, we used the Search Tool for the Retrieval of Interacting Genes/Proteins (STRING) database [17] against the 16 Mtb-dependent eQTL genes with strong interaction defined at a combined score > 700. We also conducted hypergeometric mean enrichment pathway analysis using the Molecular Signatures Database (MSigDB) C2 canonical pathways (BIOCARTA, KEGG, PID, REACTOME, WikiPathways) and C5 gene ontology biological pathways (BP) [18] with the R package clusterProfiler [94]. Pathways were significant at FDR < 0.1 and overlap > 1. GO pathways were grouped by semantic similarity > 0.85 using rrvgo.

### siRNA knockdown of selected Mtb-dependent eQTL genes

We used siRNA to silence Mtb-dependent eQTL genes, and 300 nM of two oligos per gene (siRNA identification s2032 and s2033 for *BMP6*; s3275a and s32752 for *CHMP1B*; s16450 and s16451 for *KCNK5*; s20481 and s20482 for *PIBF1*; s28111 and s28112 for *PRKAG2*; s20496 and s20497 for *TIMM44*, Life technologies) were used. As a control, Silencer Select negative control siRNAs (Silencer Select Negative Control No. 1 siRNA catalog # 4390843, Life technologies) was used at 300 nM per well. Transfection of the siRNAs into THP-1 cells was performed using Amaxa™ Cell Line Nucleofector™ Kit V (Lonza) following the manufacturer’s protocol for “Amaxa™ 4D-Nucleofector™ Protocol for THP-1 [ATCC].” After 2h, the nucleofected cells were replated in RPMI-10 (i) at a concentration of 3.0 × 10^5^ cells per well in a 24-well plate and differentiated for 24h in phorbol 12-myristate 13-acetate (PMA, Invitrogen) for DNA/RNA sensor stimulation experiments; (ii) at a concentration of 1.0 x 10^5^ cells per well in a 96-well pate and differentiated for 24h in PMA for stimulation with TLR ligands, NLRP3/NLRC4-specific proteins, as described below. Cells were washed and rested overnight in RPMI-10 without PMA at 37 °C in a humidified incubator.

siRNA knockdown experiments were repeated for *BMP6* with a 2^nd^ transfection techinque. THP-1 cells were plated at 5 x 10^5^ cells per well in a 24-well plate with RPMI-10 and PMA and rested at 37 °C in a humidified incubator. After 24h incubation, differentiated cells were washed and replated with RPMI-10 without PMA. We used 10 nM each of the two oligos (s2032 and s2033) for *BMP6* per well. As a control, Silencer Select negative control siRNAs (Silencer Select Negative Control No. 1 siRNA 4390843 and Silencer Select Negative Control No. 2 siRNA 4390846) were used at 10 nM each per well. Transfection of the pooled siRNAs into THP-1 cells was performed using Lipofectaime™ RNAiMAX transfection reagent (ThermoFisher) following the manufacturer’s protocol for “Universal Lipofectamin® RNAiMAX Reagent Protocol 2013.” After 24h of treatment with *BMP6* siRNA, cells were washed and replated with fresh RPMI-10 to each well, and treatment with DNA/RNA ligands was performed as described below.

### Cytokine expression and secretion following stimulation of cytoplasmic DNA/RNA sensing, TLR, and inflammasome pathways

For DNA/RNA sensing-specific stimulation, nucleofected THP-1 cells were treated with a medium and high dose of each DNA ligand, complexed with an equal dose of Lipofectamine 2000 for 4h. DNA ligands include: 1μg/mL and 4μg/mL of Sheared Calf Thymus DNA (Sigma Aldrich, #D1501) complexed with 1μg/mL and 4 μg/mL of Lipofectamine 2000 (Thermo Fisher Scientific); 0.1 μg/mL and 1μg/mL of Super Coiled Plasmid DNA (Invivogen) complexed with 2.5 μg/mL and 4 μg/mL of Lipofectamine 2000; 5 μM and 20 μM of cGAMP complexed with 0.4μg/mL and 4 μg/mL of Lipofectamine 2000; and 80μM cGAMP without Lipofectamine 2000. RNA ligands include: 5 μg/mL, 10μg/mL, and 20 μg/mL of poly (I:C) (Invivogen) complexed with 1 μg/mL, 2 μg/mL, and 4 μg/mL of Lipofectamine 2000, respectively. As controls, 10 ng/mL lipopolysaccharide (LPS, Invivogen); RPMI-10 complexed with an equivalent dosage of Lipofectamine 2000, RPMI-10 without Lipofectamine 2000 were used in each well. Following a 4h stimulation, cells were directly lysed in Buffer RLT Plus (Qiagen) and homogenized. RNA was isolated from lysates using the RNeasy® Plus Mini Kit (Qiagen) following the manufacturer’s recommendations. cDNA synthesis was performed using the High Capacity cDNA RT kit (Applied Biosystems). *IFNB1* and selected Mtb-dependent eQTL gene expression were measured by qRT-PCR using pre-designed TaqMan gene expression assays (Applied Biosciences).

For repeated *BMP6* knockdown experiments, RNA was isolated from lysates using the same protocol with RNeasy® Plus Mini Kit (Qiagen). Synthesis of the first strand cDNA was performed using SuperScript II reverse transcriptase and oligo (dT) primer (Invitrogen). qPCR was performed with the CFX96 real-time system (Bio-Rad) using the SsoFast EvaGreen Supermix with the LOW ROX kit (Bio-Rad). The following primers designed from PrimerBank were used. The PrimerBank identifications are *BMP6* (133930782c1) and *IFNB1* (50593016c1).

BMP6 forward: AGCGACACCACAAAGAGTTCA

BMP6 reverse: GCTGATGCTCCTGTAAGACTTGA

IFNB1 forward: ATGACCAACAAGTGTCTCCTCC

IFNB1 reverse: GGAATCCAAGCAAGTTGTAGCTC

The expression of *GAPDH* was used to standardize the samples. For knockdown efficiency analysis, mRNA levels of siRNA-treated cells were normalized to control siRNA-treated cells using the 2−ΔΔCT (cycle threshold) method to calculate fold induction. For DNA/RNA sensing-specific stimulation, the results were expressed as the normalized ratio of each expression relative to the RPMI-10 with an equivalent dosage of Lipofectamine 2000 control.

For TLR-specific stimulation, nucleofected THP-1 cells were treated with LPS (100ng/mL TLR4, Invivogen), Pam2CSK4 (0.25μg/mL TLR2/6, Invivogen), and Pam3CSK4 (0.25μg/mL TLR2/1, Invivogen) for 24h. To induce NLRP3-specific activation, cells were treated with nigericin (10 μm) for 4h following a 2h pre-stimulation with LPS at 0.1 ng/mL. NLRC4-specific stimulation was achieved using recombinant proteins consisting of Burkholderia thailandensis needle protein combined with the Bacillus anthracis lethal factor (generously provided by Russel Vance, University of California Berkeley) for 4h [95]. The supernatant from these TLR and inflammasome pathway experiments was harvested and analyzed for TNF-α, IL-6, and IL-1β cytokines using ELISA (R&D Systems).

## Study Approval

The current prospective cohort is part of the larger Kawempe Community Health Study conducted in Kampala, Uganda. Detailed information on the original study’s setting, recruitment procedure, informed consent, and ethical approval has been published elsewhere [11, 15, 86]. All participants provided written informed consent or witnessed verbal consent if they were illiterate. The study protocols were approved by the National AIDS Research Committee, the Uganda National Council on Science and Technology, and the Institutional Review Boards at the University Hospitals Cleveland Medical Center and the University of Washington.

## Data Availability

Access to raw transcriptomic data is available through the NCBI database of Genotypes and Phenotypes (dbGaP) Data Browser (https://www.ncbi.nlm.nih.gov/gap/) under accession 002445.v1.p1, but first must be approved by data access committees (DACs) for each study site (see Supplemental Methods [14]). All R code is available at https://github.com/hawn-lab/RSTR_eQTL_public.

## Acknowledgements

We thank the individual study participants of the Kawempe Community Health Study, study coordinators and the clinical and research staff including LaShaunda Malone, Keith Chervenak, Marla Manning, Dr. Mary Nsereko, Dr. Moses Joloba, Hussein Kisingo, Sophie Nalukwago, Dorcas Lamunu, Deborah Nsamba, Annet Kawuma, Saidah Menya, Joan Nassuna, Joy Beseke, Michael Odie, Henry Kawoya, Shannon Pavsek, Dr. E. Chandler Church, Anna Duewiger and Dr. Bonnie Thiel.

## Author Contributions

TRH, WHB, HMK, and CMS devised the project, the main conceptual ideas and proof outline. Clinical cohorts were established in Uganda with initial epidemiologic characterization by CMS, HMK, and WHB. JDS and GJP performed transcriptional experiments. Computational transcriptional analyses were performed by HH and KD. Statistical genetic analyses were conducted by HH, PB and CMS. Statistical descriptive analyses were conducted by HH. HH, KD and TRH drafted the manuscript with input from all authors. All authors reviewed and approved the final version of the manuscript.

