## Supplementary material for "*Mycobacterium tuberculosis*-dependent Monocyte Expression Quantitative Trait Loci and Tuberculosis Pathogenesis": Figure S1

**Figure S1. Expression plots for the remaining 9 Mtb-dependent eQTLs.** This figure displays the association between 9 Mtb-dependent eQTLs and the expression of their target genes, which are not presented in the main text. The x-axis represents the different genotypes, while the y-axis shows the normalized log2 expression of target genes. The lines indicate a linear fit derived from an additive regression model that includes an interaction term (Mtb_infection:Genotype), adjusting for age and sex. The FDR represents the significance level for eQTLs in either the Mtb-uninfected media condition or the Mtb-infected condition, not the interaction model.

ALT= alternative allele, REF= reference allele
