## Supplementary material for "*Mycobacterium tuberculosis*-dependent Monocyte Expression Quantitative Trait Loci and Tuberculosis Pathogenesis": Figure S2

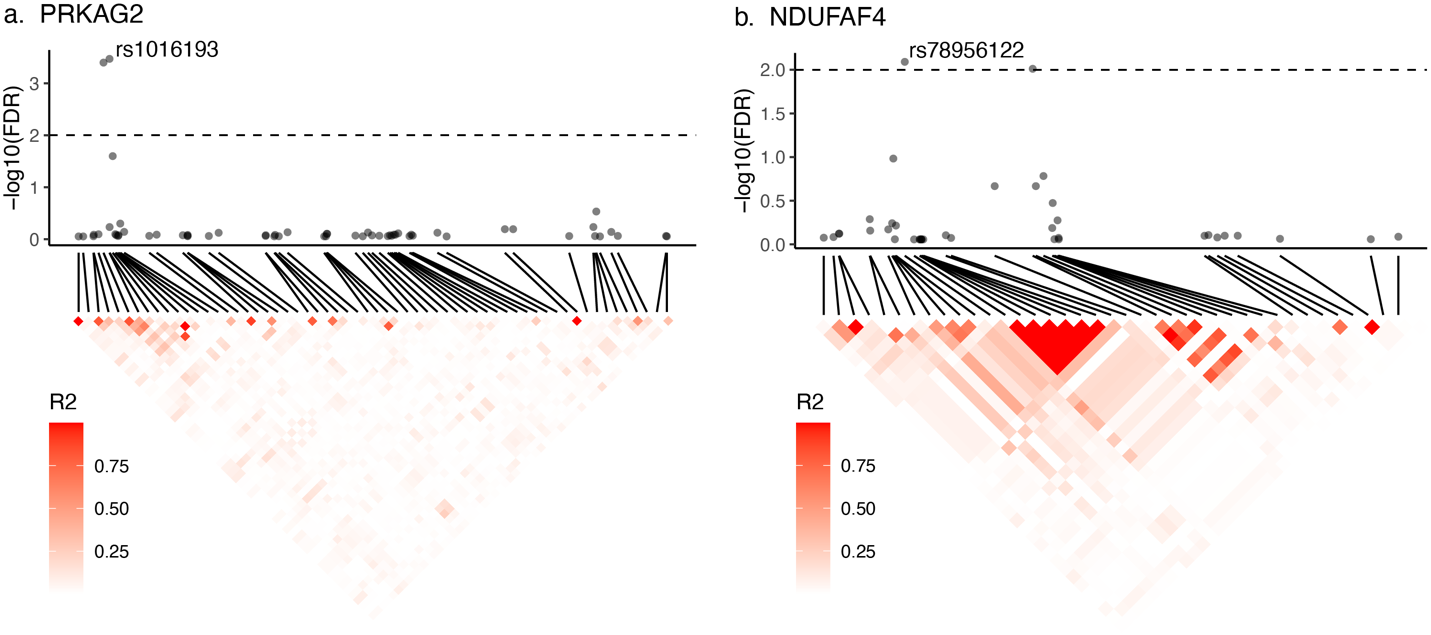

**Figure S2. Pattern of linkage disequilibrium.** PRKAG2 and NDUFAF4 had 2 cis eQTLs in high linkage disequilibrium (LD). (top) eQTL significance for SNPs within 1 MB of (a) PRKAG2 and (b) NDUFAF4. X-axis indicates chromosome position of the eQTLs. Horizontal dashed line indicates FDR = 0.01 and the lead SNP is labeled. (bottom) Heatmap indicating R^2^ LD for SNPs in this region.
