## Supplementary material for "*Mycobacterium tuberculosis*-dependent Monocyte Expression Quantitative Trait Loci and Tuberculosis Pathogenesis": Figure S3

a.

b.

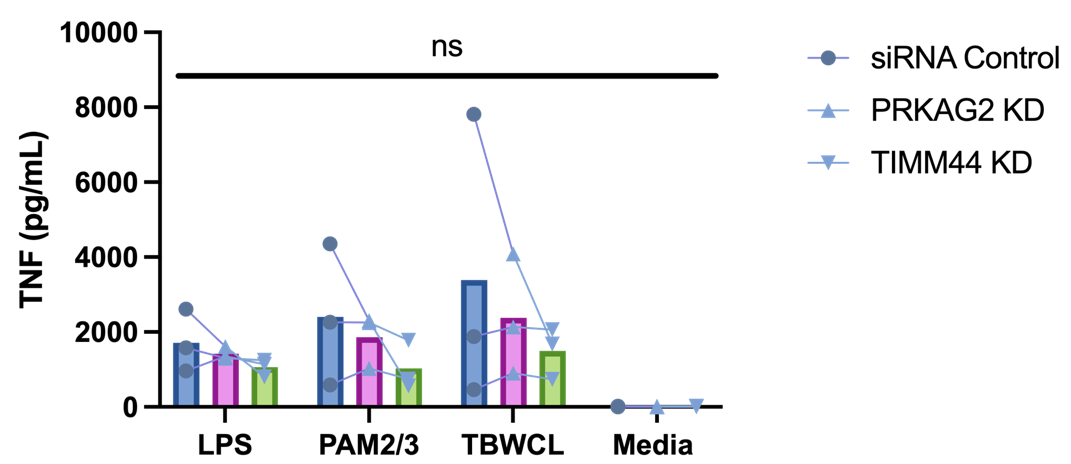

d.

c.

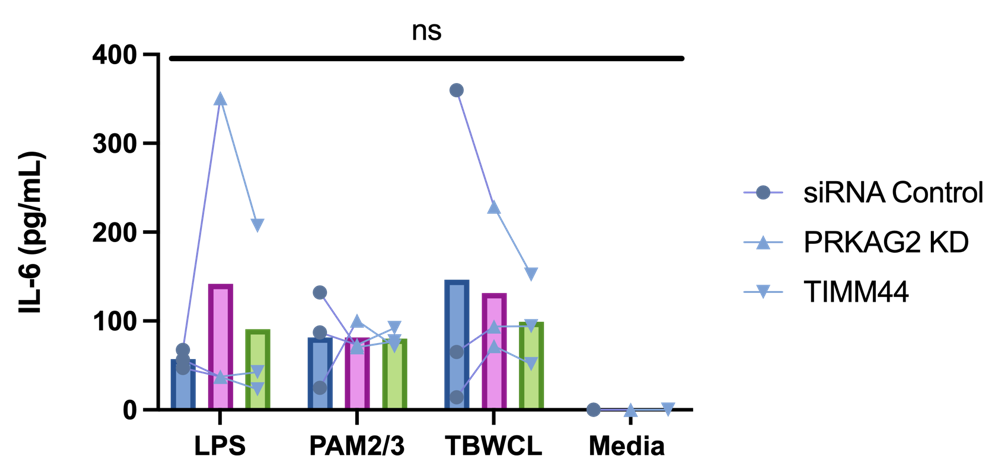

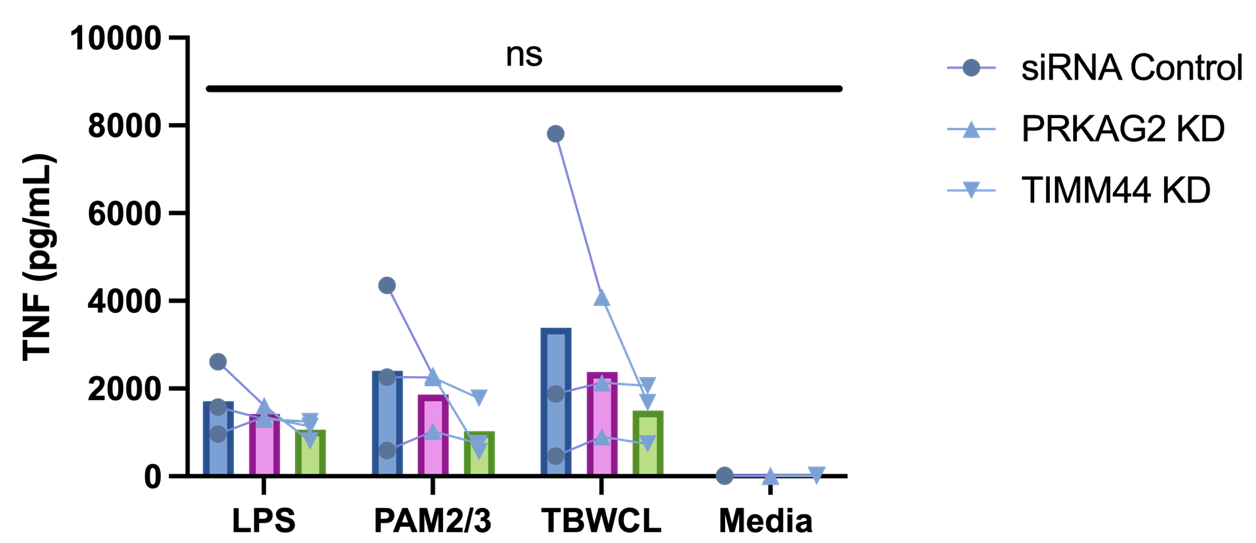

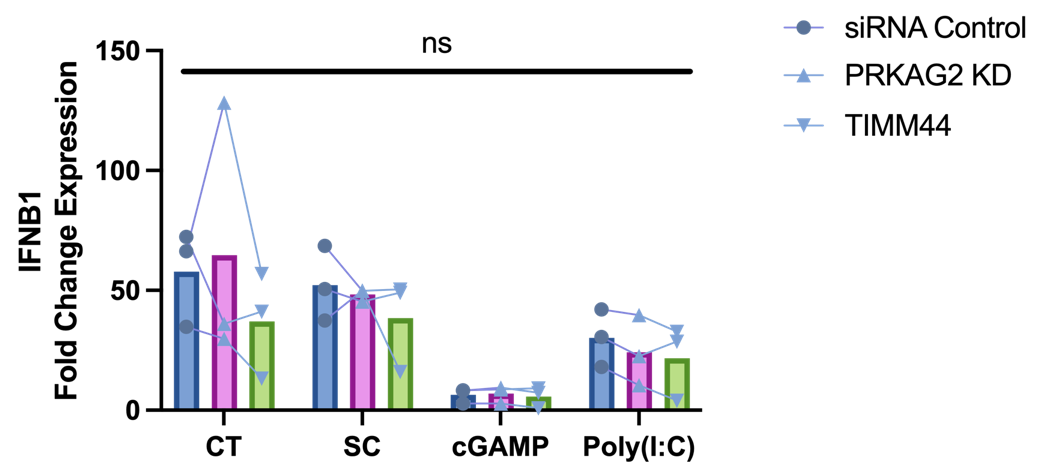

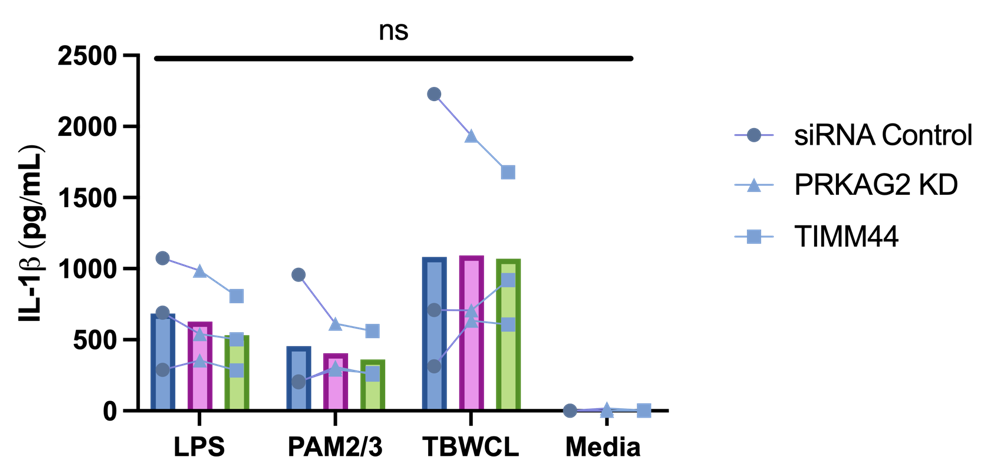

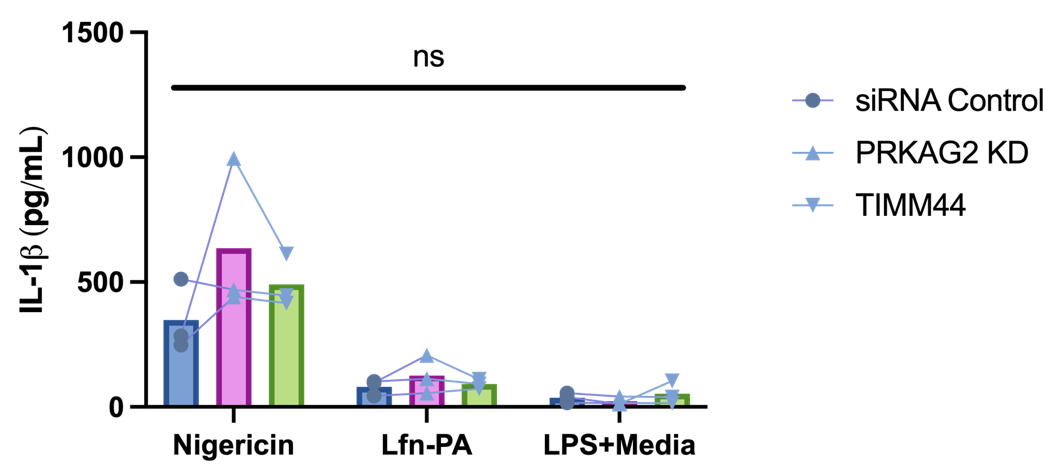

e.

**Figure S3. Cytokine responses in PRKAG2- and TIMM44-silenced THP-1 cells stimulated with DNA, TLR, and inflammasome ligands.** PRKAG2-silenced and TIMM44-silenced THP-1 cells, when stimulated with various ligands (calf thymus DNA, supercoiled plasmid DNA, cGAMP, or poly(I:C)), showed no significant changes in (a) IFNB1 expression compared to siRNA control cells. For TLR-specific stimulation, PRKAG2-silenced, TIMM44-silenced, and siRNA control THP-1 cells were stimulated with LPS, PAM2/PAM3, Mtb whole cell lysate, or media. There were no significant differences in (b) TNF, (c) IL-6, and (d) IL-1β supernatant levels at 24h stimulation. To evaluate the inflammasome-mediated IL-1β response, nucleofected cells were primed with LPS for 2h and then treated with nigericin for NLRP3-specific stimulation or *Burkholderia thailandensis* needle protein with *Bacillus anthracis* protective antigen for NLRC4-specific stimulation. There were also no significant differences observed between PRKAG2-silenced or TIMM44-silenced cells and siRNA control cells in inflammasome-mediated (e) IL-1β response. IFNB1 expression was quantified by qPCR and normalized against background induction from lipofectamine. TNF, IL-6, and IL-1β supernatant levels were measured by ELISA.

cGAMP = cyclic guanosine monophosphate-adenosine monophosphate, CT = calf thymus DNA, LPS = lipopolysaccharide, media = media control (RPMI + 10% FBS), PAM2 = Pam2CSK4, PAM3 = Pam3CSK, poly(I:C) = polyinosinic acid-polycytidylic acid, SC = supercoiled plasmid DNA, TBWCL = Mtb whole cell lysates, TLR = toll-like receptor.

ns: not significant.
