## Supplementary material for "*Mycobacterium tuberculosis*-dependent Monocyte Expression Quantitative Trait Loci and Tuberculosis Pathogenesis": Figure S4

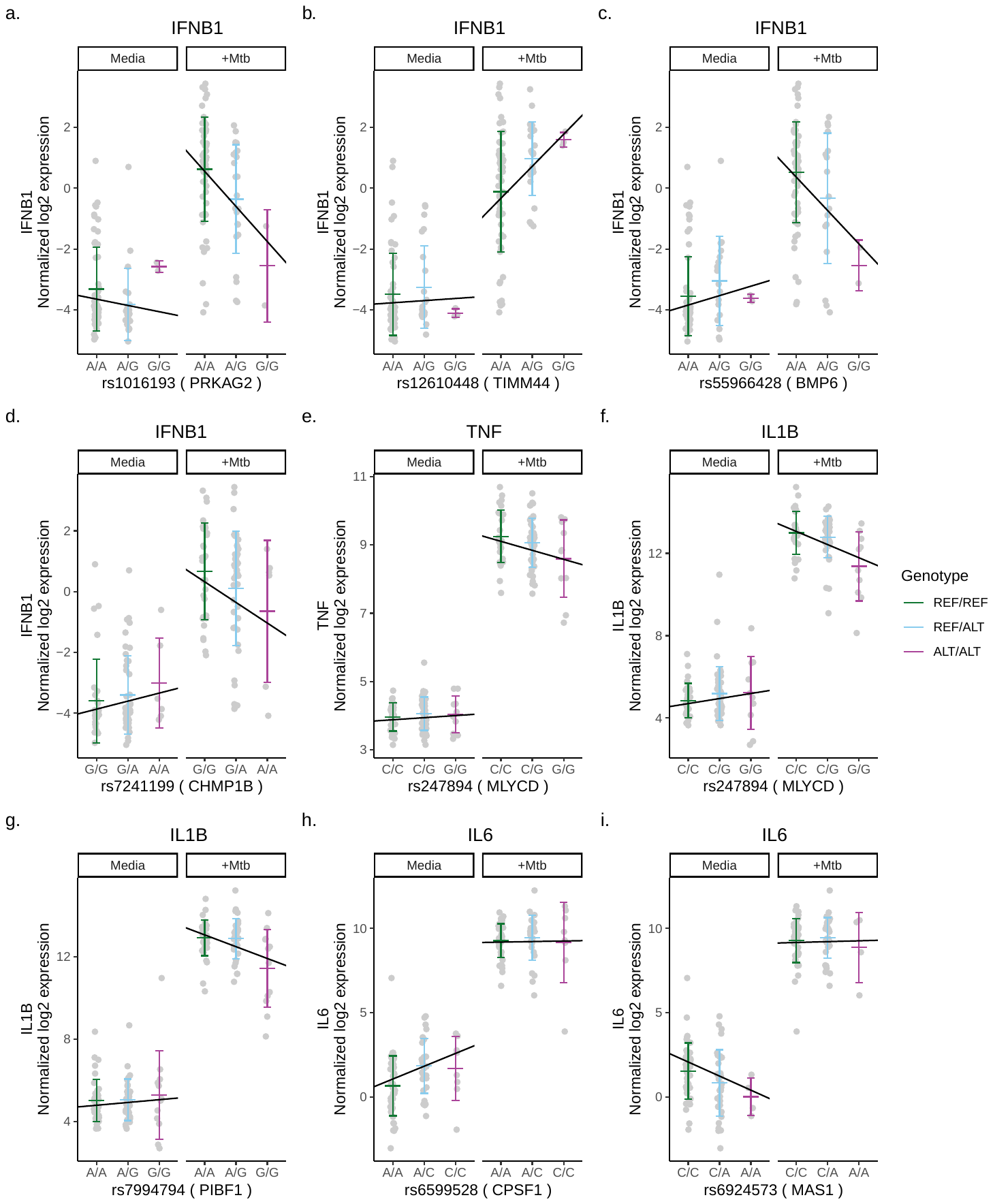

**Figure S4. Mtb-dependent eQTLs associated with Mtb-induced cytokine expression in Mtb-infected and uninfected monocytes.** While the relationship between host genotype and differences in cytokine expression was established through the subtraction of cytokine expression from Mtb-infected to uninfected conditions (i.e., normalized log2 [Mtb-infected – uninfected relative cytokine expression]), this figure depicts the correlation between Mtb-dependent eQTLs and the expression of cytokine genes in Mtb-uninfected media and Mtb-infected conditions separately. The x-axis represents different genotypes, while the y-axis represents the normalized log2 expression of cytokine genes. The slope of the lines indicates the ratio of the effect estimate to the standard error derived from a linear regression model, which estimates the relationship between the number of minor alleles and the differences in cytokine expression in each condition, controlling for age and sex (p < 0.05).

ALT= alternative allele, REF= reference allele
